# Adiposity and risk of prostate cancer death: a prospective analysis in UK Biobank and meta-analysis of published studies

**DOI:** 10.1101/2021.10.05.21264556

**Authors:** Aurora Perez-Cornago, Yashvee Dunneram, Eleanor L. Watts, Timothy J. Key, Ruth C. Travis

**Affiliations:** Cancer Epidemiology Unit, Nuffield Department of Population Health, University of Oxford, Oxford, UK

## Abstract

**Introduction:** The association of adiposity with prostate cancer specific mortality remains unclear. We examined how adiposity and its distribution relates to fatal prostate cancer by analysing data from UK Biobank, and conducting a dose-response meta-analysis to integrate existing prospective evidence. We also described the cross-sectional associations in UK Biobank of commonly used adiposity measurements with indices of adiposity estimated by imaging.

**Methods:** 218,246 men from UK Biobank who were free from cancer at baseline were included and participants were followed-up via linkage to health administrative datasets. Body mass index (BMI), total body fat percentage (using bioimpedance), waist circumference (WC) and waist-to-hip ratio (WHR) were collected at recruitment. Risk of dying from prostate cancer (primary cause) by the different adiposity measurements was estimated using multivariable-adjusted Cox proportional hazards models. Results from this and other prospective cohort studies were combined in a dose-response meta-analysis.

**Results:** In UK Biobank, 631 men died from prostate cancer over a mean follow-up of 11.5 years. The hazard ratios (HR) for prostate cancer death were 1.10 (95% confidence interval=1.00-1.21) per 5 kg/m^2^ higher BMI, 1.03 (0.96-1.11) per 5% increase in total body fat percentage, 1.09 (1.02-1.18) per 10 cm increase in WC, and 1.09 (1.02-1.16) per 0.05 increase in WHR. Our meta-analyses of prospective studies included 22,106 prostate cancer deaths for BMI, 642 for body fat percentage, 3,153 for WC and 1,611 for WHR, and the combined HRs for dying from prostate cancer for the increments above were 1.10 (1.08-1.13), 1.03 (0.96-1.11), 1.08 (1.04-1.12), and 1.07 (1.02-1.12), respectively. In up to 4,800 UK Biobank participants with magnetic resonance imaging and dual-energy X-ray absorptiometry, BMI and WC were strongly associated with imaging estimations of total and central adiposity (e.g. visceral fat, trunk fat), with associations marginally larger for WC. There might be ∼1000 fewer prostate cancer deaths per year in the UK if the mean BMI in men was reduced by 5 kg/m^2^.

**Conclusion:** Overall, we found that men with higher total and central adiposity had similarly higher risks of prostate cancer death, which may be biologically driven or due to differences in detection. In either case, these findings provide further reasons for men to maintain a healthy body weight.

## Introduction

Prostate cancer is the second most common cause of cancer death in men in the UK [1]. Age, black ethnicity, family history of prostate cancer, genetic factors, and endogenous hormones are known risk factors for prostate cancer, but apart from hormones none of them is modifiable [2-5]. While many prostate cancer tumours are indolent (slow□growing tumours), others are lethal, and these tumours may have different risk factors [3]. However, the aetiology of lethal prostate cancer is not well understood, and there is a need to identify risk factors for this clinically relevant form of the disease.

There is some evidence that relates adiposity to prostate cancer risk, but the association appears to vary by tumour subtype. Previous studies have found an inverse association of obesity with overall prostate cancer and non-aggressive forms of the disease, probably due to later diagnosis in men with obesity. However, a positive association with aggressive prostate cancer, including risk of dying from prostate cancer, has been reported [2, 6], although it is unclear whether this positive association is due to late detection (and thus more advanced tumours with poorer prognosis), a role of excessive adiposity in promoting metabolic and hormonal dysfunction that in turn may stimulate the growth and progression of prostate cancer cells [7], or a combination of both. Moreover, some evidence suggests that fat located within the abdominal cavity may be more aetiologically important for aggressive prostate cancer than total adiposity [6], and the use of “gold standard” methods to determine body fat distribution (e.g. magnetic resonance imaging (MRI)) [8, 9] may help to better understand these associations. However, due to the limited number of prostate cancer deaths in most prospective studies relatively few studies have investigated whether adiposity or its distribution is related to prostate cancer mortality [10-14], and more research is needed.

To provide reliable epidemiological evidence on the prospective association of total adiposity and its distribution with prostate cancer-specific mortality, we first report results from a prospective analysis using UK Biobank data, and then from a dose-response meta-analysis of findings from all published prospective studies. To inform the interpretation of our findings, we also describe the cross-sectional associations in UK Biobank of commonly used indices of adiposity with MRI- and dual-energy X-ray absorptiometry (DXA)-derived estimates of adiposity.

## Methods

### UK Biobank

#### Study design and population

UK Biobank is a prospective study of ∼500,000 UK adults aged 40-69 years at recruitment (including 229,000 men) established between 2006 and 2010 to study risk factors for disease. Details of the study protocol and information about data access are available online (http://www.ukbiobank.ac.uk/wp-content/uploads/2011/11/UK-Biobank-Protocol.pdf) and elsewhere [15]. All individuals provided informed consent to participate and the study was approved by the National Information Governance Board for Health and Social Care and the National Health Service North West Multicentre Research Ethics Committee (reference number 06/MRE08/65). In brief, approximately 9.2 million people living within reasonable traveling distance (∼25 km) of 1 of the 22 assessment centres across England, Wales, and Scotland were identified from National Health Service (NHS) registers and invited to participate in the study, with a participation rate of 5.5% [16].

After excluding 9,869 men with prevalent cancer (except C44: non-melanoma skin cancer), 1 man censored on entry day, and 999 men with no adiposity measurements, the analyses included a total of 218,246 men (Supplemental Figure 1).

#### Assessment of adiposity and other predictor variables

At recruitment, participants provided detailed information on a range of sociodemographic, physical, lifestyle, and health-related factors via self-completed touch-screen questionnaires and a computer assisted personal interview [16]. Anthropometric measurements (standing height, weight, waist and hip circumferences) were taken by trained research clinic staff at the assessment centre, while body mass index (BMI) was calculated and percentage body fat was estimated through bioimpedance measures [17].

#### UK Biobank imaging sub-cohort

In 2014, the UK Biobank imaging study re-invited a subsample of participants to undergo abdominal MRI and DXA, which has been detailed elsewhere [18, 19]. In brief, participants were scanned in a Siemens MAGNETOM Aera 1.5□T MRI scanner (Siemens Healthineers, Germany) using a 6-min dual-echo Dixon Vibe protocol, providing water-and-fat-separated volumetric information for fat and muscle. Body composition analyses for MRI images were performed using AMRA Profiler Research (AMRA Medical AB, Linköping, Sweden). DXA captures whole-body composition (e.g. bone, fat and lean mass) with no extensive additional processing and analysis. However, it is not possible to obtain direct compartmental volumetric measurements using this method, and therefore regional volume estimates are obtained indirectly using anatomical models [18, 19]. By January 2021, imaging data on ∼4,800 men were available. BMI, WC, hip circumference and body fat percentage were also re-assessed at the imaging visit.

#### Ascertainment of prostate cancer mortality

Our endpoint was prostate cancer as the underlying cause of death recorded on the death certificate (International Classification of Diseases Tenth revision codes: C61 [20]). Men were followed-up until 31^st^ December 2020 for England and Scotland and 19^th^ July 2020 for Wales. Mortality data were provided by NHS Digital for England and Wales and by the NHS Central Register and Information and Statistics Division for Scotland.

#### Statistical analysis in UK Biobank

### Cross-sectional analyses of adiposity measurements

Pearson correlations between different anthropometric measurements were calculated. A subsample of men had both commonly used anthropometric measures of adiposity (i.e. BMI, body fat percentage, WC and WHR) and adiposity information (MRI and DXA) from the imaging visit. Men were categorized into tenths of BMI, body fat percentage, WC and WHR, and multivariable linear regressions (adjusted for age and height) were conducted to calculate mean values from MRI and DXA. Moreover, multivariable linear regression adjusted for categories of age and height was used to estimate the mean differences in each MRI- and DXA-derived measure of body composition per 1-SD difference in the levels of each commonly used anthropometric measure of adiposity.

### Prospective analyses

Cox proportional hazards models were used to calculate hazard ratios (HRs) and 95% confidence intervals (CIs) for prostate cancer death, using age as the underlying time variable. Person-years were calculated from the date of recruitment to the date of death, loss to follow-up, or the censoring date, whichever occurred first. The proportional hazards assumption was examined using time-varying covariates and Schoenfeld residuals, and revealed no evidence of deviation. Men were categorized into fourths of adiposity measurements based on the distribution in the cohort. We also modelled HRs per predefined increments and categories of the adiposity measurements: (i) BMI [per 5 kg/m^2^ increase, and as predefined World Health Organization (WHO) categories [21] (<25, 25–29.9, and ≥30 kg/m^2^)]; (ii) body fat percentage (per 5% increase); (iii) WC [per 10 cm increase, and as predefined WHO categories [22] (<94, 94-101.9, ≥ 102 cm)]; and (iv) WHR [per 0.05 unit increase, and as predefined WHO categories [22] (<0.90, ≥ 0.90)]. Potential nonlinear associations between the anthropometric variables and prostate cancer mortality were evaluated using likelihood ratio tests comparing the model with the anthropometric variable entered as an ordered categorical (ordinal) variable to a model with the categorical variable treated as continuous, and no evidence of non-linearity was observed.

Adjustment covariates were defined *a priori* based on previous analyses by our group using UK Biobank data [23]. The minimally-adjusted models were stratified by geographical region of recruitment (ten UK regions) and age (<45, 45–49, 50–54, 55–59, 60–64, ≥65 years) at recruitment. The fully adjusted model was further adjusted for Townsend deprivation score (fifths, unknown [0.1%]), ethnic group (white, mixed background, Asian, black, other, and unknown [0.6%]), height (<170, 170–174.9, 175–179.9, ≥180 cm, and unknown [0.2%]), lives with a wife or partner (no, yes), cigarette smoking (never, former, current 1-<15 cigarettes per day, current ≥15 cigarettes per day, current but number of cigarettes per day unknown, and smoking status unknown [0.6%]), physical activity (low [0–9.9 METs/week], moderate [10–49.9 METs/week], and high [≥50 METs/week], unknown [3.6%]), alcohol consumption (non-drinkers, <1–9.9, 10–19.9, ≥20 g ethanol/day, unknown [0.5%]), and diabetes (no, yes, and unknown [0.5%]).

### Sensitivity analyses

We also performed the following sensitivity analyses: excluding the first 5 years of follow-up; excluding men with BMI ≥27.5 kg/m^2^; excluding men with BMI ≥25 kg/m^2^; excluding extreme values (percentiles outside 1-99); excluding men <50 years of age at recruitment; running the statistical analyses per 1 standard deviation (SD) increment, using the BMI-adjusted residuals of WC (or WHR, depending on which one is the exposure of interest) by regressing these variables in a linear regression model and using the residuals (that are statistically independent BMI) as the exposures of interest.

#### Dose-response meta-analysis

We searched on PubMed, Embase, and Web of Science for prospective studies examining the association of BMI, WC and WHR with prostate cancer as the underlying cause of death, independently by two researchers up to 15th March (please see details in the **Supplementary Methods** and Supplementary Figure 2). We excluded reviews, abstract-only publications or editorials.

In the dose-response meta-analysis, we calculated the HR estimates in the studies that reported results for a different increment (e.g. per 1 SD increase) or from the categorical data using generalized least-squares[24] for the increments mentioned above before pooling the data from the different prospective studies (details in **Supplementary Methods**). After that, we pooled study-specific log HRs to obtain a summarized effect size using a fixed effects model. The I^2^ statistic was used to assess heterogeneity across studies.

#### Population-attributable risk

The number of prostate cancer deaths attributable to obesity (population-attributable risk (PAR)) in the UK was calculated by applying estimates of relative risk from our dose response meta-analyses and information on the prevalence of obesity in English men aged 55-64 years (mean BMI 28.9 kg/m^2^) [25].

All analyses were performed using Stata version 14.1 (Stata Corporation, College Station, TX, USA), and figures were plotted in R version 3.2.3. All tests of significance were two-sided, and P-values <0.05 were considered statistically significant.

## Results

### UK Biobank participants’ characteristics

After an average of 11.5 years of follow-up, a total of 631 (0.3%) men died from prostate cancer among the 218,246 men included in the UK Biobank study. The main baseline characteristics of the participants are shown in **Table 1**, while baseline characteristics of participants according to categories of BMI and WC are reported in Supplementary Tables 4 and 5. 12.4% of men reported that they were current cigarette smokers, 43.3% reported drinking ≥20 g of alcohol per day and 27.6% of men reported being physically inactive. Men who died from prostate cancer had higher values of all adiposity measurements at recruitment (**Table 1**). Moreover, men with higher adiposity at baseline were more likely to be older, drink ≥20 g of alcohol per day, be physically inactive, and to have hypertension and diabetes than men in the lowest quartiles of BMI and WC (Supplementary Tables 4 and 5).

**Table 1:** baseline characteristics in all men and in men who died from prostate cancer in men from UK Biobank.

| Characteristics at baseline | All men | Men who died from prostate cancer |
| --- | --- | --- |
| No. of men | 218246 | 631 |
| **Sociodemographic** |  |  |
| Age at recruitment (years), mean (SD) | 56.5 (8.2) | 63.3 (4.7) |
| Most deprived quintile, n (%) | 44,807 (20.5) | 110 (17.4) |
| No qualifications, n (%) | 29,466 (13.5) | 81 (12.8) |
| Black ethnicity, n (%) | 3,225 (1.5) | 4 (0.6) |
| Not in paid/self-employment, % (n) | 84,580 (38.8) | 406 (64.3) |
| Living with partner, n (%) | 166,386 (76.2) | 479 (75.9) |
| **Anthropometric** |  |  |
| Height (cm), mean (SD) | 175.6 (6.8) | 175.3 (7.0) |
| BMI (kg/m<sup>2</sup>), mean (SD) | 27.8 (4.3) | 28.2 (4.4) |
| Body fat (%), mean (SD) | 25.3 (5.8) | 26.2 (5.8) |
| Waist circumference (cm), mean (SD) | 96.9 (11.4) | 98.9 (11.3) |
| Waist to hip ratio, mean (SD) | 0.936 (0.065) | 0.950 (0.065) |
| **Lifestyle** |  |  |
| Current cigarette smokers, n (%) | 27,249 (12.4) | 66 (9.9) |
| Drinking alcohol $\geq 20$ g/day, n (%) | 94,408 (43.3) | 285 (45.2) |
| Physically inactive, n (%) | 60,230 (27.6) | 165 (26.1) |
| **Health status** |  |  |
| Vasectomy, n (%) | 11,344 (5.2) | 24 (3.8) |
| Hypertension, n (%) | 113,878 (52.2) | 398 (61.9) |
| Diabetes, n (%) | 15,088 (6.9) | 63 (10.0) |
| **Prostate specific factors prior recruitment** |  |  |
| PSA test, n (%) | 60,442 (27.7) | 196 (31.1) |
| Enlarged prostate, n (%) | 7,074 (3.2) | 34 (5.4) |
| Family history of prostate cancer, n (%) | 16,383 (7.5) | 60 (9.5) |

Abbreviations: BMI, body mass index; PSA, prostate specific antigen.

### Cross-sectional associations in UK Biobank

BMI, body fat percentage and WC were strongly correlated (correlation coefficients (*r*) = 0.79-0.88), although these measures were less strongly correlated with WHR (*r* = 0.59-0.79, Supplementary Table 7). BMI and WC were strongly associated with total and central adiposity (e.g. visceral fat, trunk fat) obtained from MRI- and DXA-derived measures of body composition. The magnitudes of the associations with MRI- and DXA-measures of body fat were generally marginally larger for WC, while the associations with WHR were somewhat smaller (**Table 2**, Supplementary Tables 8-11). Muscle fat mass infiltration and liver proton density fat fraction were not strongly correlated with the commonly used anthropometric measurements (*r* = 0.36-0.48) (Supplementary Tables 8 & 9).

**Table 2.** Mean difference in MRI- and DXA-derived body fat compartments per 1 SD higher levels of BMI, body fat percentage, waist circumference, and waist to hip ratio in men from UK Biobank.

| Adiposity measure | Mean difference per 1 SD increase (95% CI) |  |  |  |
| --- | --- | --- | --- | --- |
|  | BMI | Body fat % | Waist circumference | WHR |
| **MRI (max n=4,631)** |  |  |  |  |
| Total adipose tissue volume, L | 6.16 (6.05-6.27) | 5.73 (5.61-5.84) | 6.62 (6.51-6.74) | 4.61 (4.42-4.81) |
| Visceral adipose tissue, L | 1.86 (1.81-1.90) | 1.80 (1.76-1.84) | 2.00 (1.96-2.05) | 1.75 (1.69-1.81) |
| Abdominal subcutaneous adipose tissue volume, L | 2.21 (2.17-2.25) | 2.03 (1.98-2.08) | 2.35 (2.31-2.39) | 1.62 (1.55-1.69) |
| Muscle fat infiltration, % | 0.77 (0.72-0.81) | 0.93 (0.88-0.97) | 0.90 (0.85-0.95) | 0.81 (0.76-0.86) |
| Liver proton density fat fraction, % | 2.10 (1.91-2.29) | 2.04 (1.85-2.24) | 2.13 (1.92-2.33) | 2.01 (1.79-2.23) |
| **DXA (max n=2,361)** |  |  |  |  |
| Total tissue fat % | 4.97 (4.80-5.14) | 6.00 (5.87-6.13) | 5.64 (5.47-5.82) | 4.79 (4.55-5.03) |
| Trunk fat mass | 7.16 (7.08-7.24) | 6.24 (6.05-6.43) | 7.27 (7.12-7.41) | 5.55 (5.27-5.82) |
| Trunk fat % | 6.60 (6.38-6.83) | 7.78 (7.60-7.96) | 7.48 (7.25-7.70) | 6.63 (6.31-6.94) |
| Android fat mass | 1,112 (1,090-1,134) | 1,109 (1,082-1,137) | 1,196 (1,171-1,220) | 979 (935-1,023) |
| Android fat % | 7.54 (7.26-7.81) | 8.95 (8.72-9.17) | 8.54 (8.26-8.82) | 7.68 (7.31-8.05) |
| Gynoid fat mass | 1,129 (1,104-1,155) | 1,103 (1,071-1,135) | 1,183 (1,152-1,213) | 787 (735-840) |
| Gynoid fat % | 4.27 (4.10-4.44) | 5.13 (4.98-5.28) | 4.86 (4.69-5.04) | 3.72 (3.48-3.97) |
| VAT mass | 809 (786-832) | 771 (745-797) | 863 (837-888) | 744 (709-779) |
| VAT % | 858 (833-882) | 817 (790-845) | 914 (887-941) | 789 (752-825) |

Multivariable linear regression model adjusted by age and height.
Abbreviations: BMI, body mass index; DXA, dual-energy x-ray absorptiometry; MRI, magnetic resonance imaging; VAT, visceral adipose tissue; WHR, waist to hip ratio.

### Prospective analysis in UK Biobank

The multivariable-adjusted associations of BMI, body fat percentage, WC and WHR with prostate cancer mortality are reported in **Figure 1** (minimally-adjusted associations are shown in Supplementary Table 12). There were no large changes between the minimally- and the multivariable-adjusted models. The HR per 5 kg/m^2^ higher BMI was 1.10 (95% CI 1.00-1.21, P-trend=0.059), although when we compared the highest quartile with the lowest, BMI was not significantly associated with prostate cancer death (HR=1.04, 0.83-1.31). Total body fat percentage was not significantly associated with prostate cancer death (HR per 5% increase=1.03, 0.96-1.11), whereas WC (HR per 10 cm increase=1.09, 1.02-1.18) and WHR (HR per 0.05 increase=1.09, 1.02-1.16) were significantly associated with risk of dying from prostate cancer; when the highest quartiles were compared to the lowest the HRs were 1.36 (1.07-1.72) for WC and 1.28 (1.01-1.62) for WHR. BMI, WC and WHR were also categorised according to the WHO cut-off points, and we found that those with a WC ≥ 102 cm had a higher risk of dying from prostate cancer compared to those with lower WC (<94cm) (1.22, 1.00-1.48), while there were no significant associations with BMI and WHR when comparing categories defined with the WHO cut-points (Supplementary Table 13).

**Figure 1.**
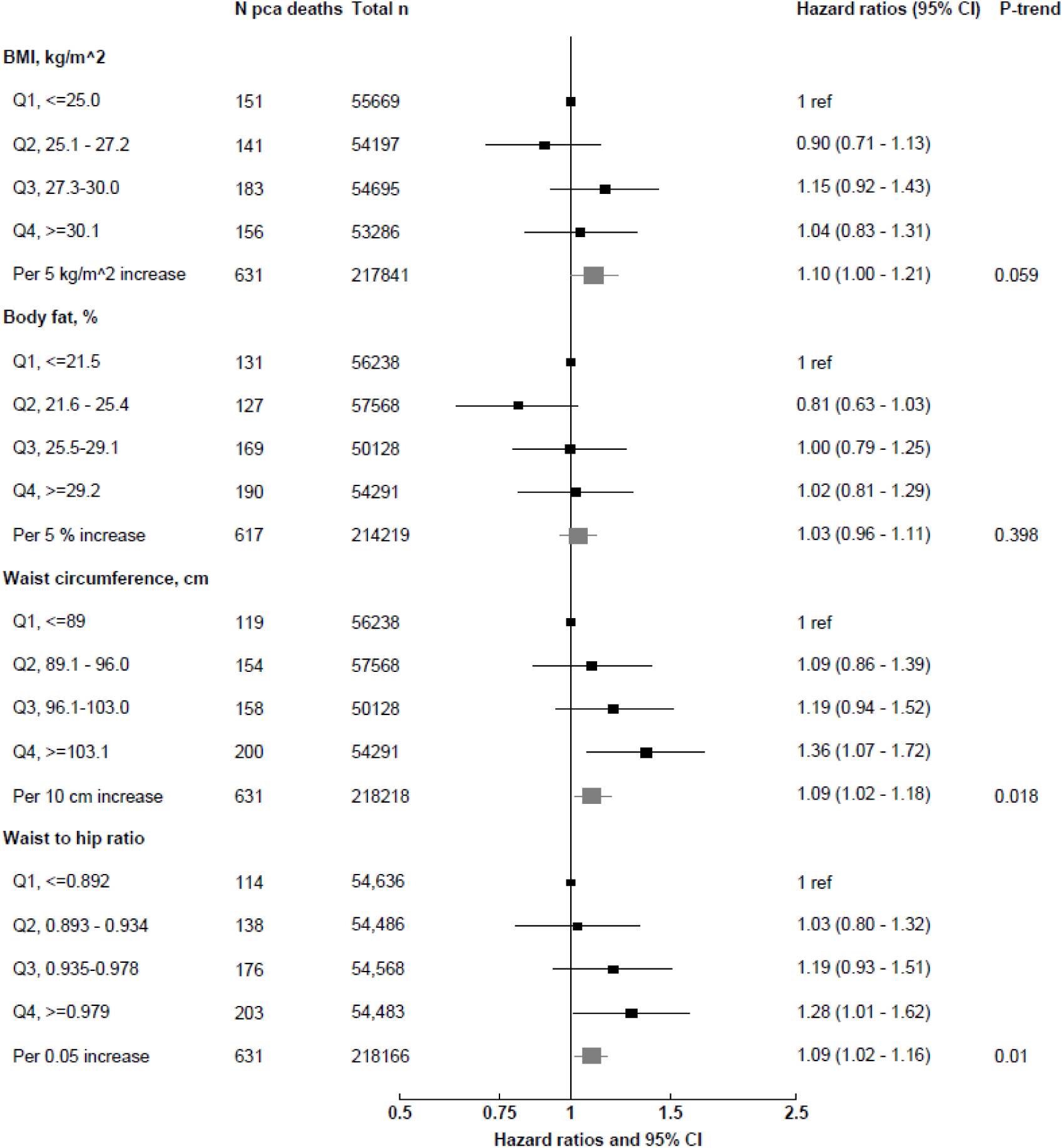
Multivariable-adjusted hazard ratios (95% CI) for prostate cancer death in relation to adiposity measurements at baseline in men from UK Biobank. Abbreviations: BMI, body mass index. Cox regression analyses. All models are stratified by region and age at recruitment and adjusted for age (underlying time variable), Townsend deprivation score, ethnicity, lives with a wife or partner, smoking, physical activity, alcohol consumption, height, and diabetes. Full details for each covariate are provided in the statistical section.

In sensitivity analyses, we found that the associations remained largely unchanged after excluding the first 5 years of follow-up (**Table 3**). When the associations between commonly used anthropometric measurements and prostate cancer death were assessed using a 1 SD increment of each exposure of interest (4.3 kg/m^2^ for BMI, 5.8% for body fat percentage, 11.4 cm for WC, and 0.065 for WHR), the HRs were 1.08 (1.00-1.18), 1.04 (0.95-1.13), 1.11 (1.02-1.20), and 1.11 (1.03-1.21), respectively for BMI, body fat percentage, WC, and WHR (Table 3). After excluding men with BMI ≥25 kg/m^2^, we found that the magnitude of the association of WHR with risk of death from prostate cancer became larger (1.19 [1.01-1.40]), although with wider confidence intervals (Table 3).

**Table 3.** Sensitivity analyses. Multivariable-adjusted hazard ratios (95 % CI) for prostate cancer death in relation to adiposity measurements at recruitment in 218,246 men from UK Biobank.

|  | BMI |  |  | Body fat percentage |  |  | Waist circumference |  |  | Waist to hip ratio |  |  |
| --- | --- | --- | --- | --- | --- | --- | --- | --- | --- | --- | --- | --- |
|  | n cases | Per 5 kg/m<sup>2</sup> increase | p-trend<sup>1</sup> | n cases | Per 5 % increase | p-trend<sup>1</sup> | n cases | Per 10 cm increase | p-trend<sup>1</sup> | n cases | Per 0.05 unit increase | p-trend<sup>1</sup> |
| Overall | 631 | 1.10 (1.00-1.21) | 0.059 | 617 | 1.03 (0.96-1.11) | 0.398 | 631 | 1.09 (1.02-1.18) | 0.018 | 631 | 1.09 (1.02-1.16) | 0.010 |
| Excluding first 5 years of follow-up | 537 | 1.10 (0.99-1.22) | 0.076 | 529 | 1.04 (0.96-1.13) | 0.324 | 537 | 1.09 (1.00-1.18) | 0.040 | 537 | 1.08 (1.01-1.15) | 0.035 |
| Excluding men with BMI $\geq 25$ kg/m<sup>2</sup>, per 1 SD increment | 147 | 0.99 (0.84-1.18) | 0.938 | 143 | 0.91 (0.77-1.08) | 0.283 | 147 | 1.13 (0.95-1.35) | 0.167 | 147 | 1.19 (1.01-1.40) | 0.035 |
| Excluding extreme values: percentiles 1-99 | 631 | 1.10 (1.00-1.21) | 0.059 | 617 | 1.03 (0.96-1.11) | 0.398 | 631 | 1.09 (1.02-1.18) | 0.018 | 631 | 1.09 (1.02-1.16) | 0.010 |
| Excluding men <50 years of age | 625 | 1.10 (0.99-1.21) | 0.065 | 611 | 1.03 (0.95-1.11) | 0.467 | 625 | 1.09 (1.01-1.17) | 0.024 | 625 | 1.08 (1.02-1.15) | 0.013 |
| Per 1 SD increment | 631 | 1.08 (1.00-1.18) | 0.059 | 617 | 1.04 (0.95-1.13) | 0.398 | 631 | 1.11 (1.02-1.20) | 0.018 | 631 | 1.11 (1.03-1.21) | 0.010 |
| Residuals<sup>2</sup> |  |  |  |  |  |  | 631 | 1.14 (0.97-1.34) | 0.117 | 631 | 1.07 (1.00-1.16) | 0.073 |

Abbreviations: BMI, body mass index.
<sup>1</sup> *P*-values for trend are obtained by entering the anthropometric variable per increment in the Cox regression model.
<sup>2</sup> HR (95% CI) is from the multiple adjusted model (above) after accounting for the residuals of waist circumference and waist to hip ratio regressed on BMI for analyses of waist circumference and waist to hip ratio as exposures.

When we used BMI-adjusted residuals of WC and WHR as exposures the associations of both WC and WHR with risk of prostate cancer death became larger (**Table 3**).

### Dose-response meta-analyses

A total of 19, 1, 6, and 3 prospective studies (in addition to the current report on UK Biobank) were identified that had reported on BMI, body fat percentage, WC, and/or WHR, respectively, in relation to prostate cancer-specific mortality (Supplementary Tables 1-3). When these results were combined with UK Biobank, data from a total of 22,106 (for BMI), 642 (for body fat percentage), 3,153 (for WC), 1,611 (for WHR) men who died from prostate cancer were available.

In the dose-response meta-analyses, the weighted average HRs were 1.10 (1.08-1.13) for every 5 kg/m^2^ increase in BMI, 1.03 (0.96-1.11) for every 5% increase in body fat percentage, 1.08 (1.04-1.12) for every 10 cm increase in WC, and 1.07 (1.02-1.12) for every 0.05 increase in WHR. There was no statistically significant heterogeneity between studies for any of these associations (**Figures 2-4**).

**Figure 2.**
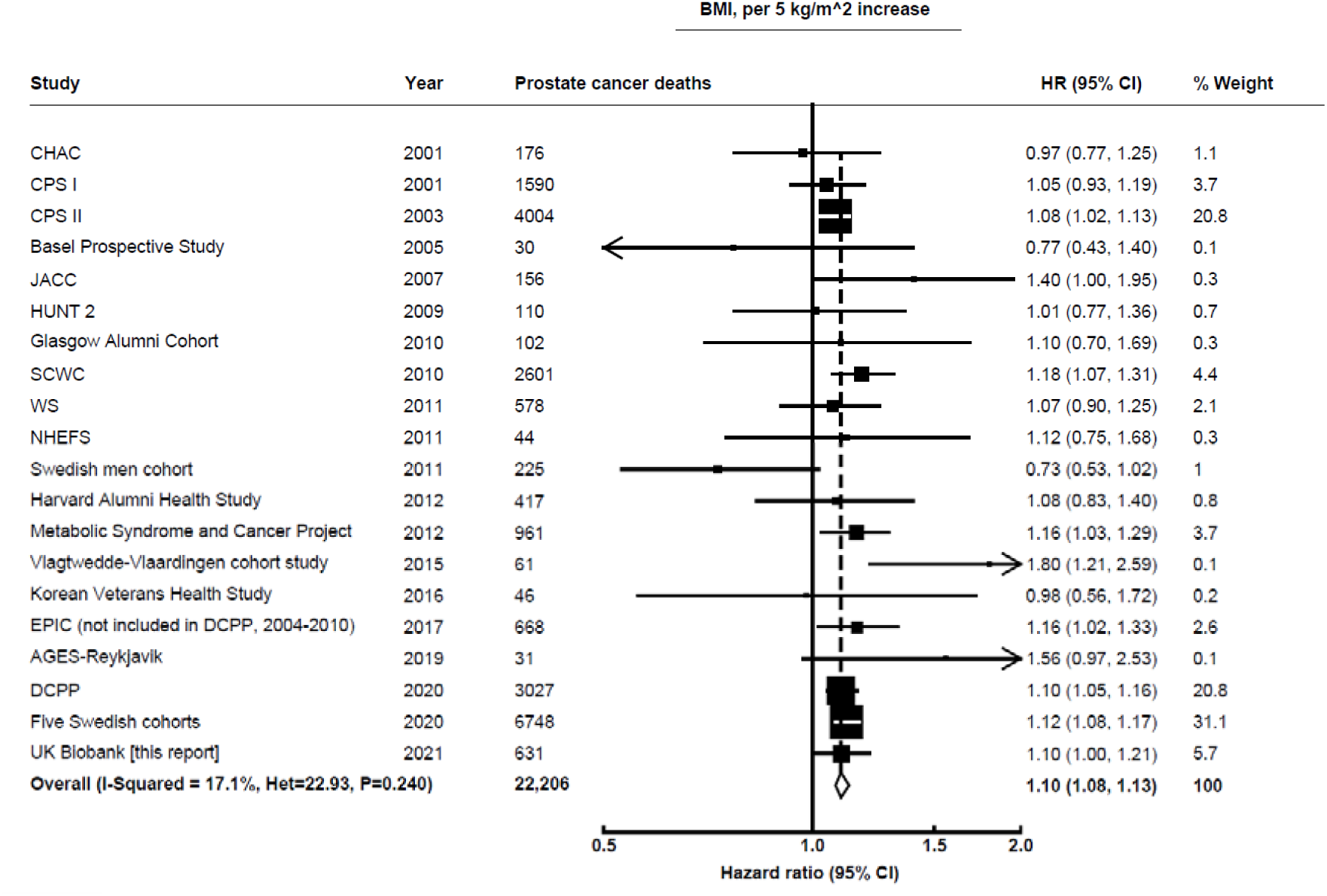
Meta-analysis of prospective studies on the risk of prostate cancer death in relation to BMI. Study-specific hazard ratios (HR) are represented by squares (with their 95% confidence intervals [CIs] as lines). HRs were combined using inverse-variance-weighted averages of the log HRs in the separate studies, yielding a result and its 95% CI, which is plotted as a diamond. Please see Supplementary Table 1 for further details about each study. Abbreviations: AGES-Reykjavik, Age, Gene/Environment Susceptibility-Reykjavik; CHAC, The Chicago Heart Association; CPS I, Cancer Prevention Study I Nutrition Cohort Study; CPS II, Cancer Prevention Study II Nutrition Cohort Study; DCPP, Diet and Cancer Pooling Project; EPIC, European Prospective Investigation into Cancer and Nutrition; JACC, Japan Collaborative Cohort Study; HUNT 2, Nord-Trøndelag Health Study; NHEFS, Nutrition Examination Survey Epidemiology Follow-Up Study; SCWC, Swedish Construction Workers Cohort; WS, Whitehall study.

**Figure 3.**
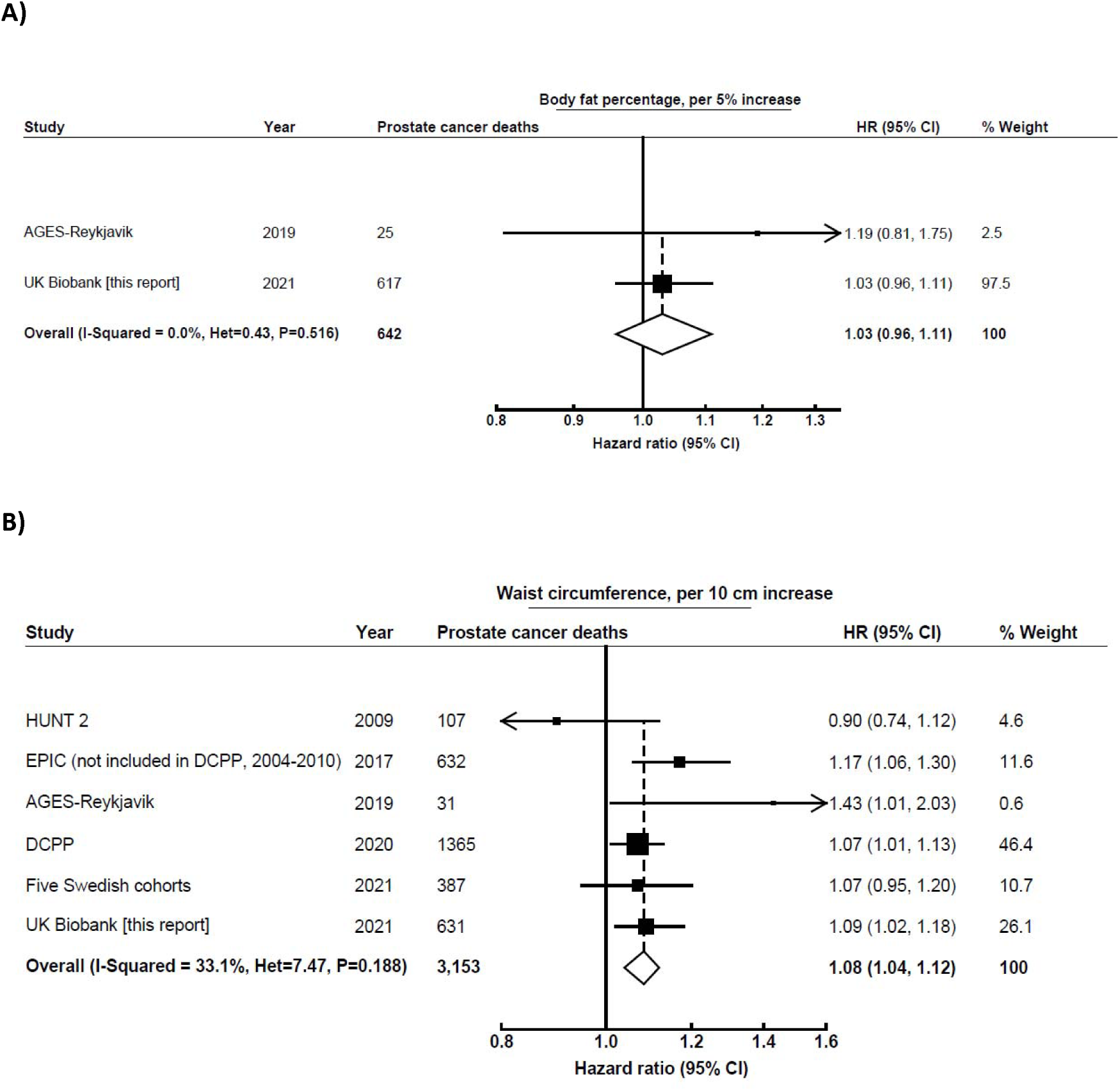
Meta-analysis of prospective studies on the risk of prostate cancer death in relation to body fat percentage (A) and waist circumference (B). Study-specific hazard ratios (HR) are represented by squares (with their 95% confidence intervals [CIs] as lines). HRs were combined using inverse-variance-weighted averages of the log HRs in the separate studies, yielding a result and its 95% CI, which is plotted as a diamond. Please see Supplementary Table 2 for further details about each study. Abbreviations: AGES-Reykjavik, Age, Gene/Environment Susceptibility-Reykjavik; DCPP, Diet and Cancer Pooling Project; EPIC, European Prospective Investigation into Cancer and Nutrition; HUNT 2, Nord-Trøndelag Health Study.

**Figure 4.**
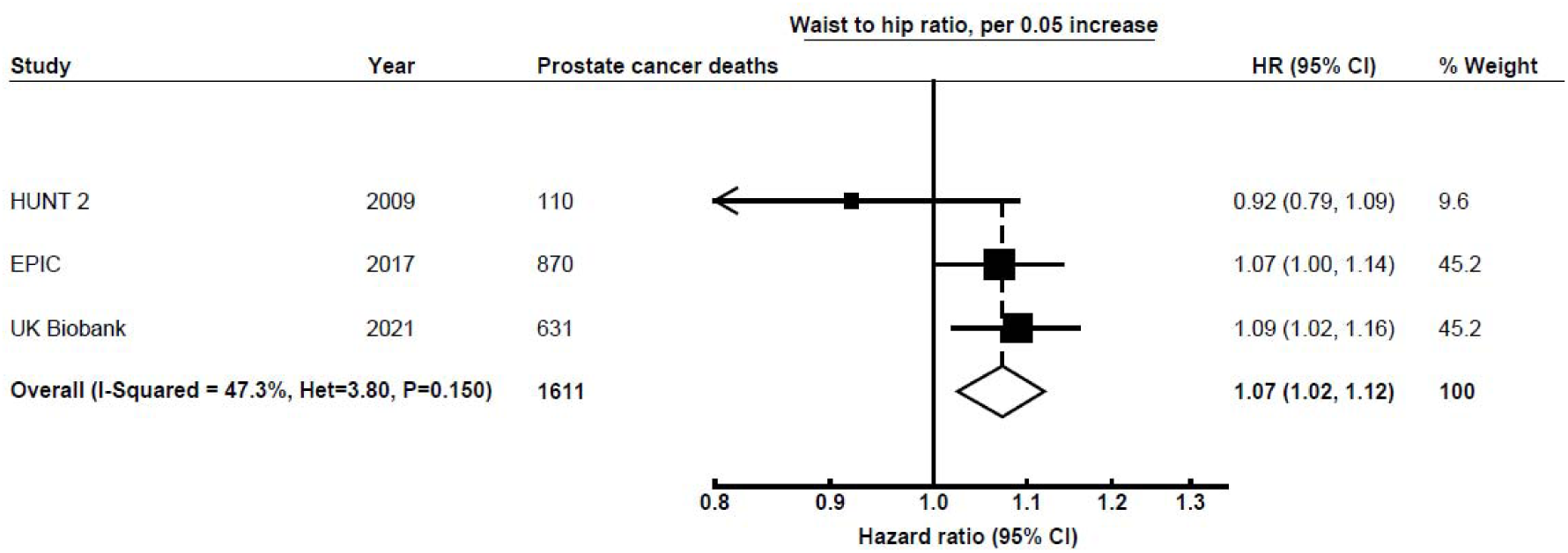
Meta-analysis of prospective studies on the risk of prostate cancer death in relation to waist to hip ratio. Study-specific hazard ratios (HR) are represented by squares (with their 95% confidence intervals [CIs] as lines). HRs were combined using inverse-variance-weighted averages of the log HRs in the separate studies, yielding a result and its 95% CI, which is plotted as a diamond. Please see Supplementary Table 3 for further details about each study. Abbreviations: EPIC, European Prospective Investigation into Cancer and Nutrition; HUNT 2, Nord-Trøndelag Health Study.

A total of 11,900 men die from prostate cancer each year in the UK [26]. If it is assumed that the HR from our meta-analysis is unbiased and that in the English general population in men aged 55-64 years the mean BMI is 28.9 kg/m^2^, a reduction of 5 kg/m^2^ would decrease mean BMI to within the ideal BMI range and would lead to about 1,000 less prostate cancer deaths annually in the UK.

## Discussion

Findings from new analyses in the UK Biobank cohort and from a dose-response meta-analysis combining results from the UK Biobank and other prospective studies showed positive associations for both total and central adiposity in relation to prostate cancer death. In a subsample of UK Biobank participants with MRI- and DXA-derived estimates of body adiposity, we found that BMI and WC were strongly associated with total and central adiposity (e.g. visceral fat, trunk fat) from imaging data, with associations marginally larger for WC, whereas the associations of WHR with the MRI and DXA estimations were smaller.

Our dose-response meta-analysis included more than double the number of prostate cancer deaths than previous meta-analyses, and it suggested similar associations with prostate cancer mortality for total and central adiposity. The prospective data on central adiposity as assessed by WC and WHR in relation to subsequent prostate cancer death, however, are still relatively limited (6 studies with a total of 3,153 prostate cancer deaths) [6, 27-30], and more studies are needed to confirm the magnitude of the association with central adiposity. The latest World Cancer Research Fund (WCRF) meta-analysis on prostate cancer published in 2014 also reported a positive association between overall adiposity (assessed using BMI) and prostate cancer death based on 10,100 prostate cancer deaths in a total of 12 studies; however, it did not have enough data from previous prospective studies to look at the association with central adiposity measurements (i.e. WC and WHR) [2]. A more recent pooled analysis of individual participant data from up to 15 prospective studies that included 3,000 prostate cancer deaths for total adiposity and 1,300 for central adiposity found a positive association of both total and central adiposity with prostate cancer mortality [27].

Obesity is defined as excessive fat accumulation, but some commonly used measures of adiposity such as BMI do not differentiate reliably between fat and fat-free mass. WC has been proposed as a better marker than BMI of adiposity in middle-aged men [31]; however, in men in UK Biobank WC and BMI are highly correlated and they showed similar associations with “gold standard” measurements of adiposity (MRI and DXA) in our cross-sectional analyses. We found that both BMI and WC were strongly positively associated with total and central adiposity (e.g. visceral fat, trunk fat) from the imaging data, with associations marginally larger for WC, whereas the associations were smaller for WHR. Previous studies have suggested that visceral fat is more strongly related than subcutaneous fat to metabolic and hormonal dysfunction (e.g. insulin resistance, impaired glucose metabolism, low-grade inflammation) [32, 33], and hence might play a more important role in prostate cancer progression. To the best of our knowledge, only one small prospective study (n<2000 men, 31 prostate cancer deaths) has examined the associations between different fat depots (visceral and subcutaneous fat, and thigh intermuscular and subcutaneous fat), using computed tomography scans and risk of prostate cancer death, finding similar positive associations of specific fat depots measured by CT, BMI and WC with aggressive and fatal prostate cancer [29]; due to the small sample size of this study and the lack of other prospective studies looking at fat depots as exposures, more research looking at body fat distribution based on imaging data in relation to risk of prostate cancer mortality is needed before conclusions can be drawn.

Additionally, commonly used measures of adiposity also do not assess ectopic fat (fat stored in tissues other than adipose tissue, for example liver proton density fat fraction and muscle mass infiltration) [31]. Correspondingly, in UK Biobank we found that while associations of BMI and WC with visceral fat estimates from imaging data were large, the associations of BMI and WC with liver proton density fat fraction and muscle mass infiltration were weaker. The weaker associations and also the biological plausibility of associations with liver fat suggest that there may be additional utility in assessing the associations with risk of prostate cancer mortality using these measures.

Obesity has been associated with a higher risk of being diagnosed with high grade prostate tumours [27], which have poorer prognosis, and several biological mechanisms have been proposed for the association between adiposity and prostate cancer development and progression [7]. However, it does not seem likely that any of the known biological risk factors for prostate cancer may mediate this association. Both IGF-I and free testosterone are positively associated with prostate cancer risk in observational and Mendelian randomization studies [4, 5], however, men with obesity have moderately lower concentrations of IGF-I and free testosterone than men with a healthy BMI [34, 35]. Higher BMI is associated with lower concentrations of IGFBP□1 and IGFBP□2 [34], which might lead to higher bioavailability of IGF-I, but evidence on the association of these binding proteins with prostate cancer mortality is limited. Other biomarkers that are altered in men with obesity include pro-inflammatory cytokines [36] and oxidative stress biomarkers [37], which have been hypothesised to increase prostate cancer risk [38, 39]. Further, some evidence suggests that visceral, periprostatic and pelvic fat might promote proliferation and inhibit apoptosis of prostate cancer cells through paracrine mechanisms, such as the secretion of growth factors and pro-inflammatory cytokines [40-42]. However, although these mechanisms are possible, the current evidence is too limited to suggest that they mediate the positive association between adiposity and prostate cancer mortality and more research is needed. Emerging tools such as metabolomics, proteomics, and epigenetics and the integration of this information with the gold-standard measures of adiposity have the potential to reveal novel mechanisms through which adiposity may increase prostate cancer development and progression [43].

Although the association between adiposity and prostate cancer mortality may be mediated by metabolic changes, it is likely that differences in detection also play a role. Men with obesity may have a delayed diagnosis of prostate cancer tumours due to their lower prostate-specific antigen (PSA) concentrations (owing to increased blood volume with higher BMI) and to the greater difficulty of performing a thorough digital rectal examination, and thus their lower likelihood of undergoing a biopsy [44-46]. For example, a previous meta-analysis showed that compared to men with a normal weight, those with obesity have on average 12.9% lower PSA concentrations [45]. Furthermore, enlarged prostates may make cancer detection by biopsy more difficult, due to the large size also resulting in a higher likelihood of the needles missing the cancer [44-46]. A later detection of a prostate tumour will lead to worse prognosis and a higher risk of dying from the disease. Some studies have investigated the association of classic measurements of adiposity with prostate cancer mortality by stage of the disease [44] and further research about how obesity impacts the pathway to prostate cancer diagnosis is needed.

Strengths of our analyses in UK Biobank include its prospective design, detailed information on potential confounders, and the large sample size. Analyses excluding the first 5 years of follow-up did not suggest that the observed associations were influenced by reverse causality, but substantially longer follow-up time is needed to be more confident about this. Adiposity measurements were assessed by trained research clinic staff instead of being self-reported, and we had high-quality body composition data (i.e. DXA- and MRI-derived adiposity measurements) in a subsample, which allowed us to assess the associations of commonly used adiposity measurements with “gold standard” measurements.

Our analyses also have some limitations. UK Biobank includes participants from multiple regions across the UK, including deprived areas; however, it may suffer from selection bias as it is not representative of the whole UK population [16, 47], although the directions of some major risk factor associations in the UK Biobank seem to be generalizable [48]. As in every observational study, residual confounding is possible in both our prospective analysis in UK Biobank and the meta-analysis. Moreover, there may be some misclassification of the underlying cause of death, which could be differential; obese men with prostate cancer are at increased risk of dying from several conditions, and some may die for example from cardiovascular disease but have their cause of death recorded as prostate cancer. Finally, due to the small number of prostate cancer deaths (probably due to the limited follow-up time, as 78% survive prostate cancer after ≥10 years [1]), we may have had limited power to find associations with overall adiposity (i.e. BMI and body fat percentage) in UK Biobank; we also had limited data in our meta-analysis for central adiposity.

In summary, the totality of prospective evidence indicates that men with higher adiposity (both total and central adiposity) have a higher risk of dying from prostate cancer than men with a healthy weight. Prospective studies using high quality measurements of adiposity distribution (e.g. MRI measurements) and with more data on stage, grade, and clinical information on disease progression, together with better understanding of the biological pathways, are needed to disentangle whether the association is biologically driven or due to differences in detection, but in either case, these findings provide further reason for men to maintain a healthy body weight.

## Data Availability

The datasets generated/and or analysed in the current study will be made available for bona fide researchers who apply to use the UK Biobank data set by registering and applying at http://www.ukbiobank.ac.uk/register-apply.

## Acknowledgements

This research has been conducted using the UK Biobank Resource under application numbers 24494. We would like to thank Georgina K. Fensom for her contributions to the preparation and standardization. We also thank all participants, researchers, and support staff who made the study possible.

## Abbreviations

(BMI): Body Mass Index
(CIs): confidence intervals
(DXA): dual-energy X-ray absorptiometry
(EPIC): European Prospective Investigation into Cancer and Nutrition
(IARC): International Agency for Research on Cancer
(ICD): International Statistical Classification of Diseases
(IGF-I): insulin-like growth factor-I
(HRs): hazard ratios
(MRI): magnetic resonance imaging
(NHS): National Health Service
(PCa): prostate cancer
(PSA): prostate-specific antigen
(UK): United Kingdom
(WC): waist circumference
(WCRF): World Cancer Research Fund
(WHR): waist to hip ratio

## Conflict of Interest

None disclosed.

## Funding

This research was funded by Cancer Research UK Population Research Fellowship number C60192/A28516 and Cancer Research UK programme grant number C8221/A29017. APC is also supported by the World Cancer Research Fund (WCRF UK), as part of the Word Cancer Research Fund International grant programme (2019/1953). ELW is supported by the NDPH Early Career Research Fellowship.

## Notes

### Competing Interest Statement

The authors have declared no competing interest.

### Author Declarations

All individuals provided informed consent to participate and the study was approved by the National Information Governance Board for Health and Social Care and the National Health Service North West Multicentre Research Ethics Committee (reference number 06/MRE08/65).

