## Supplementary material for "Adiposity and risk of prostate cancer death: a prospective analysis in UK Biobank and meta-analysis of published studies"

### Supplementary materials

### Supplementary methods

*Meta-analyses from prospective studies*

The latest World Cancer Research Fund (WCRF) meta-analysis did not have enough data from previous prospective studies to look at the association of central adiposity measurements (i.e. waist circumference and waist to hip ratio (WHR)) with prostate cancer mortality [1]. Moreover, a recent pooled analysis from prospective studies found a positive association of both total and central adiposity with prostate cancer mortality, but this study included 22% of all the worldwide prospective data for total adiposity and 50% for central adiposity [2].

To put our findings in UK Biobank into the context of previous research, we conducted an updated meta-analysis combining our results with those from previously published prospective studies of the association between total and central adiposity and prostate cancer death, following standard criteria for meta-analyses (the MOOSE guidelines) [3].

Literature search

We searched on PubMed, Embase, and Web of Science for prospective studies examining the association of BMI, waist circumference and WHR with prostate cancer as the underlying cause of death independently by two researchers up to 15th March 2021. We used the search terms “obesity”, “adiposity”, “body mass index”, “body mass”, “waist circumference”, “waist hip ratio”, “anthropometry”, “body composition”, “body size”, “body fat”, “prospective”, “cohort”, “prostate”, “cancer”, “carcinoma”, “tumour”, “neoplasm”, “death”, “mortality”, “fatal”, and “lethal”. After the removal of duplicate studies, titles and abstracts were independently screened by the two trained researchers using the Rayyan QRCI web application (https://rayyan.qcri.org/) [4].

Study selection

The inclusion criteria for the studies were: 1) prospective cohort studies; 2) studies that investigated the associations between BMI, body fat, waist circumference and/or WHR and prostate cancer as the underlying cause of death; 3) studies reporting hazard ratio (HR) or relative risk (RR) with the corresponding measure of variability [95% confidence intervals (CI)].

When the same cohort study published more than one original articles looking at these associations, the paper reporting the longest follow-up time was kept. Since the previous pooled analysis of individual participant data from prospective studies did not report risk of prostate cancer mortality separately in the individual studies and most of these studies have not published an original article looking at adiposity measurements and prostate cancer death, we included the pooled estimate published in the pooling project [2], and those studies included in this pooling project were not separately included in the meta-analysis (Supplementary Figure 2).

Data extraction

For each study we extracted the fully adjusted HRs and their 95% CIs of the lineal (per increment) association between the adiposity measurement and prostate cancer mortality. If comparison per increment was not available we extracted the categorical comparison. For all studies, only results with the most comprehensive adjustment for confounders were considered.

Supplementary tables 1-3 show the name of the first author, publication year, country or region, age at recruitment, years of follow-up, sample size and number of prostate cancer deaths, confounder adjustments used in each study, comparison made, HR estimates and their corresponding 95% CI, and conversion of HR and 95% CI to per increment if per increment results were not reported in the original study.

Displaying of findings

In order to include all prospective analyses on the same continuous scale (e.g. per 5 kg/m$^2$ increase in BMI), we estimated these effect measures from the available data reported in each study as follows:

- Studies that reported associations in the same continuous scale that the one we want to use in our continuous meta-analysis: This continuous scale was used in our meta-analysis.
- Studies that reported associations in a different continuous scale that the one we want to use in our continuous meta-analysis: HR and 95% CI were rescaled to the change used in this meta-analyses by $HR_y$=$(HR_x)^{y/x}$, where the HR for an increase in $y$ units of the predictor variable ($HR_y$) is equal to the HR for an increase in $x$ units ($HR_x$) raised to the power of $y/x$; the sample principle applies for the upper and lower 95% CI.
- Studies that reported categorical associations instead of associations in a continuous scale:

For all studies, an effect estimate and its standard error (SE) on the same scale (e.g. per 5 kg/m$^2$ increase in BMI) are necessary for inclusion in a ‘continuous’ meta-analysis. For BMI-prostate cancer studies, the effect estimate could be hazard ratio (HR) and risk ratio (RR). Summary relative risks combining study-specific results were estimated by calculating the weighted average of the study-specific logarithms of the relative risks, with weights proportional to the inverses of the variances of the study-specific log relative risks. (NB: calculation of such a weighted average is sometimes referred to as a fixed-effect meta-analysis). Chi-squared tests were used to assess heterogeneity across studies.

### Supplementary tables and figures

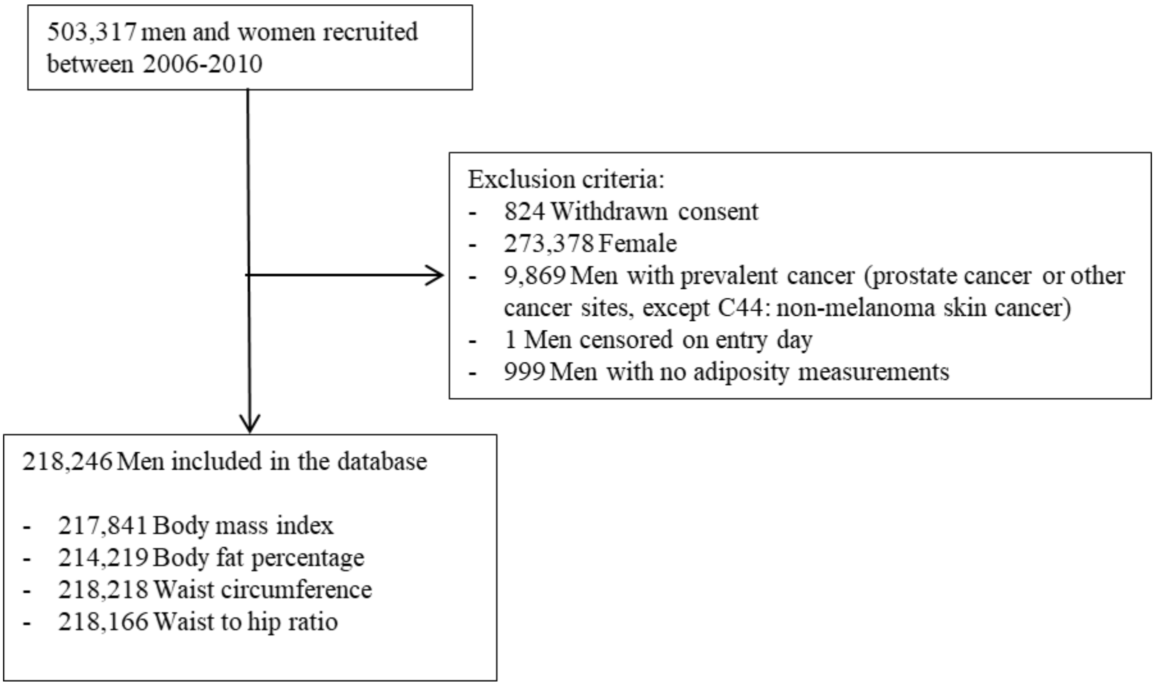

**Supplementary Figure 1.** Flow chart of the study participants in UK Biobank.

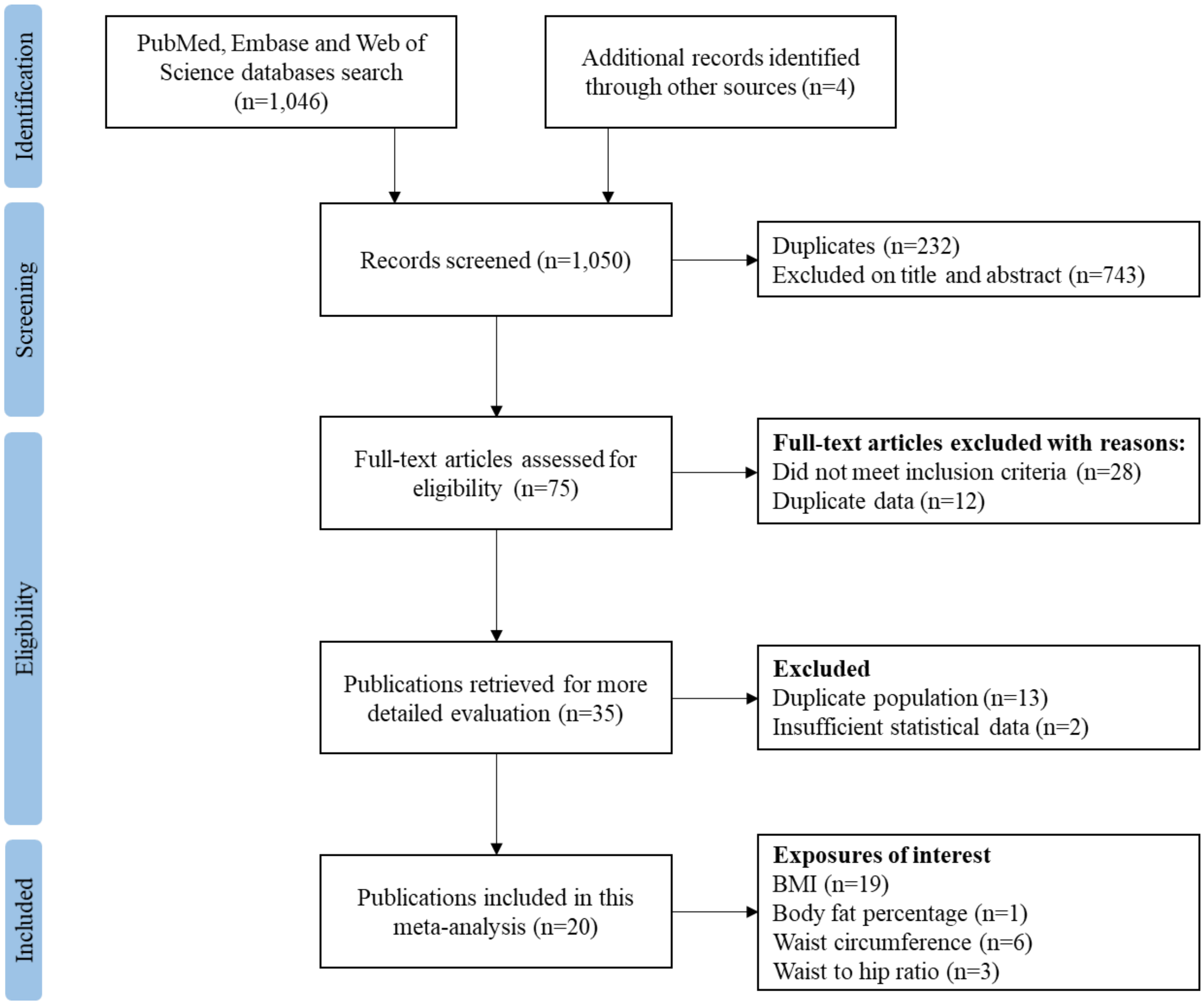

**Supplementary Figure 2.** Flow diagram of literature search and study selection for the meta-analysis.

**Supplementary table 1.** Characteristics of prospective studies and previous individual participant data meta-analysis of body mass index and prostate cancer death.

| Authors, Year (Reference) | Study | Country (Age, follow-up) | Pca death/total sample | Exposure categories | HR | 95% CI | Covariates | Conversion of HR (95% CI) to per 5 kg/m² increment [1] |
|---|---|---|---|---|---|---|---|---|
| **Gapstur et al., 2001** [5] | The Chicago Heart Association (CHAC) | USA (Mean age at baseline: 40y Mean follow-up: 27y) | 176/20,433 | Per 1 SD increase (SD=4 kg/m²) | 0.98 | 0.81-1.12 | Age, plasma glucose concentrations, heart rate, education, race | 0.97 (0.77-1.15) |
| **Rodriguez et al., 2001** [6] | Cancer Prevention Study I Nutrition Cohort Study (CPS I) | USA (Median age at baseline: 52y Mean follow-up: 13y) | 1590/456,490 | ≥32.50 kg/m² *vs* 18.50–22.49 kg/m² | 1.34 | 0.93-1.94 | Age at interview, race, height, education, exercise, smoking status, and family history of prostate cancer | 1.05 (0.93-1.19) |
| **Calle et al., 2003** [7] | Cancer Prevention Study II Nutrition Cohort Study (CPS II) | USA (Mean age at baseline: 57y Mean follow-up: 16y) | 4004/404,576 | ≥35 vs. < 25 kg/m² | 1.34 | 0.98-1.83 | Age, education, smoking status and number of cigarettes smoked, physical activity, alcohol use, marital status, race, aspirin use, fat consumption, and vegetable consumption | 1.08 (1.02-1.13) |
| **Eichholzera et al., 2005** [8] | Basel Prospective Study | Switzerland (Mean age at baseline: not available Mean follow-up: 17y) | 30/2974 | Per unit increase | 0.95 | 0.93-1.18 | Smoking status and age group | 0.77 (0.43-1.40) (final estimate taken from WCRF meta-analysis as the lower CI didn't look correct in the original publication |
| **Fujino et al., 2007** [9] | Japan Collaborative Cohort Study (JACC) | Japan (Mean age at baseline: not available Mean follow-up: ~5y (exact follow-up not given)) | 156/~110,700 (exact total n not included in original publication | ≥30 vs. 18.5-24.9 kg/m² | 0.87 | 0.12-6.29 | Age and area of study | 1.40 (1.00-1.95) |
| **Martin et al., 2009** [10] | Nord-Trøndelag Health Study | Norway (Mean age at baseline: 48y | 110/29,364 | Per 1 SD increase, 3.5 kg/m² | 1.01 | 0.83-1.24 | Age, height, smoking, marital status, education, physical activity, | 1.01 (0.77-1.36) |

| | | | | | | | | |
|---|---|---|---|---|---|---|---|---|
| | (HUNT 2) | Mean follow-up: 9.3y) | | | | | International Prostate Symptom Score | |
| **Burton et al., 2010** [11] | The Glasgow Alumni Cohort | UK (Mean age at baseline: 20y<br>Mean follow-up: 49y) | 102/9,549 | Per 1 $kg/m^2$ increase | 1.02 | 0.93-1.11 | Smoking, father's social class, and height | 1.10 (0.70-1.68) |
| **Batty et al., 2011** [12] | Whitehall study (WS) | UK (Mean age at baseline: ~50y (exact age not given)<br>Mean follow-up: 40y) | 578/17,934 | Per 1 SD increase, 2.96 $kg/m^2$ | 1.04 | 0.94-1.14 | Plasma cholesterol, physical activity, socio-economic status, diabetes/blood glucose, marital status, pulmonary function, height, age at risk, smoking, and diastolic and systolic blood pressure | 1.07 (0.90-1.25) |
| **Dehal et al., 2011** [13] | Nutrition Examination Survey Epidemiology Follow-Up Study (NHEFS) | USA (Mean age at baseline: 47y<br>Mean follow-up: 17y) | 44/3,127 | ≥30 vs. 18.5-24.9 $kg/m^2$ | 1.36 | 0.53-3.47 | Race, education attainment, family income level, marital status, types of residence area assessed at the baseline, alcohol drinking, cigarette smoking, and frequency of eating fruit and vegetables | 1.12 (0.75-1.68) |
| **Discacciati et al., 2011** [14] | Swedish men cohort | Sweden (age at baseline: aged 45–79y<br>Mean follow-up: 9y) | 225/36,959 | Per 5 $km/m^2$ increase | 0.73 | 0.53-1.02 | BMI at age 30 years, age at baseline, total energy intake, total physical activity, years of education, smoking status, family history of prostate cancer and personal history of diabetes | 0.73 (0.53-1.02) |
| **Gray et al., 2012** [15] | Harvard Alumni Health Study | USA (Mean age at baseline: 18.4y<br>Mean follow-up: 56.5y) | 417/19,593 | 2.56 $kg/m^2$ | 1.04 | 0.91–1.19 | Age, cigarette smoking status and physical activity at college entry and BMI in 1962/1966 | 1.08 (0.83-1.40) |
| **Haggstrom et al., 2012** [16] | Metabolic Syndrome and Cancer Project | USA (Mean age at baseline: 44y<br>Mean follow-up: 12y) | 961/289,866 | Quintiles, Q5 *vs* Q1 | 1.36 | 1.08-1.71 | Smoking and quintiles of BMI (except for BMI) and stratified for subcohort, 5 birth cohorts, and 5 categories of age at measurement. | 1.16 (1.03-1.29) |
| **Taghizadeh et al., 2015** [17] | Vlagtwedde-Vlaardingen cohort study | The Netherlands (Mean age at baseline: 45.9y<br>Mean follow-up: 15.5y) | 61/3718 | ≥30 vs. 18.5-24.9 $kg/m^2$ | 3.33 | 1.31-8.46 | Age, smoking habits, and place of residence | 1.8 (1.21-2.59) |
| **Hong et al., 2016** [18] | Korean Veterans Health Study | Korean (Mean age at baseline: 58.9y | 46/113,478 | Per 5 $kg/m^2$ increase | 0.98 | 0.56-1.72 | Age at baseline, smoking status, alcohol consumption, monthly | 0.98 (0.56-1.72) |

| | | | | | | | | |
|---|---|---|---|---|---|---|---|---|
| | | Mean follow-up: 6.4y) | | | | | household income, and physical activity | |
| **Perez-Cornago et al., 2017 [19]** | EPIC (not included in DCPP, follow-up from 2004-2010) | Europe (Mean age at baseline: 52y<br>Mean follow-up: 6y) | 670/129,008 | Per 5 $kg/m^2$ | 1.16 | 1.02-1.33 | Age, education level, smoking status, marital status, diabetes, and physical activity | 1.16 (1.02-1.33) |
| **Dickerman et al., 2019 [20]** | Age, Gene/Environment Susceptibility–Reykjavik (AGES-Reykjavik) study | Iceland (Mean age at baseline: not available<br>Mean follow-up: 10.4y) | 31/1832 | Per 5 $kg/m^2$ increase | 1.56 | 0.97-2.53 | Age at study entry, family history of prostate cancer, smoking status, education, physical activity, and physician visit over the past 12 months | 1.56 (0.97-2.53) |
| **Jochems et al., 2020 [21]** | Five Swedish cohorts pooling project | Sweden (Mean age at baseline: 37.5y<br>Mean follow-up: 28y) | 6748/431,902<br>(170 deaths from MDCS and 54 deaths from VIP likely to also be included in EPIC above) | Per 5 $kg/m^2$ increase | 1.12 | 1.08-1.17 | Stratified by cohort and birth period, and adjusted for baseline age, baseline smoking status, healthcare region, country of birth and education | 1.12 (1.08-1.17) |
| **Genkinger et al., 2020 [2]** | Diet and Cancer Pooling Project (DCPP) [2] | World-wide pooling project (Mean age at baseline: not available<br>Mean follow-up: not available) | 3027/830,772<br>(279 deaths from CPS-II likely to also be included in the separate CPS-II above) | Per 5 $kg/m^2$ | 1.10 | 1.05-1.16 | Age, year of questionnaire return, race, education, marital status, alcohol, smoking habits, height, physical activity, prostate cancer family history, personal history of diabetes, multiple vitamin use, dietary calcium, dietary lycopene, and total energy intake | 1.10 (1.05-1.16) |
| **Perez-Cornago et al., [this report]** | UK Biobank | UK (Mean age at baseline: 56.5y<br>Mean follow-up: 10.9y) | 571/217,841 | Per 5 $kg/m^2$ | 1.08 | 0.97-1.19 | Region, age at recruitment, Townsend deprivation score, ethnicity, lives with a wife or partner, smoking, physical activity, alcohol consumption, height, and diabetes | 1.08 (0.97-1.19) |

[1] Conversion explained in the Supplementary methods

[2] Studies included: ATBC, Alpha-Tocopherol Beta-Carotene Cancer Prevention Study; CPS II, Cancer Prevention Study II Nutrition Cohort; COSM, Cohort of Swedish Men; EPIC European Prospective Investigation into Cancer and Nutrition (follow-up only from baseline to 2004); HPFS, Health Professionals Follow-up Study; MCCS, Melbourne Collaborative Cohort Study; MDCS, Malmö Diet and Cancer Study; MEC, Multiethnic Cohort; NLCS, Netherlands Cohort Study; NIH-AARP, The NIH-AARP Diet and Health Study; and PLCO, Prostate, Lung, Colorectal, Ovarian Cancer Screening Trial; VIP, Västerbotten Intervention Project.

Abbreviations: BMI, body mass index; CI, confidence interval; HR, hazard ratio; Pca, prostate cancer.

**Supplementary table 2**. Characteristics of prospective studies and previous individual participant data meta-analysis of body fat percentage, waist circumference and prostate cancer death.

| Authors, Year (Reference) | Study | Country (Age, follow-up) | Pca death/total sample | Exposure categories | HR | 95% CI | Covariates | Conversion of HR (95% CI) to per 5 % increment [1] |
|---|---|---|---|---|---|---|---|---|
| **Body fat percentage** | | | | | | | | |
| Dickerman et al., 2019 [20] | Age, Gene/Environment Susceptibility–Reykjavik (AGES-Reykjavik) study | Iceland (Mean age at baseline: not available<br>Mean follow-up: 10.4y) | 25/1425 | Per 1 SD increase, 5.3 % | 1.20 | 0.80-1.81 | Age at study entry, family history of prostate cancer, smoking status, education, physical activity, and physician visit over the past 12 months | 1.19 (0.81-1.75) |
| **Waist circumference** | | | | | | | | **Conversion of HR (95% CI) to per 10 cm increment [1]** |
| Martin et al., 2009 [10] | Nord-Trøndelag Health Study (HUNT 2) | Norway (Mean age at baseline: 48y<br>Mean follow-up: 9.3y) | 107/29,364 | Per 1 SD increase, 9.4 cm | 0.91 | 0.74-1.12 | Age, height, smoking, marital status, education, physical activity, International Prostate Symptom Score | 0.90 (0.74-1.12) |
| Perez-Cornago et al., 2017 [19] | EPIC (not included in DCPP, follow-up from 2004-2010) | Europe (Mean age at baseline: 52y<br>Mean follow-up: 6y) | 632/129,008 | Per 10 cm increase | 1.17 | 1.06-1.30 | Age, education level, smoking status, marital status, diabetes, and physical activity | 1.17 (1.06-1.30) |
| Dickerman et al., 2019 [20] | Age, Gene/Environment Susceptibility–Reykjavik (AGES-Reykjavik) study | Iceland (Mean age at baseline: not available<br>Mean follow-up: 10.4y) | 31/1832 | Per 1 SD increase, 10.3 cm | 1.45 | 1.01 -2.07 | Age at study entry, family history of prostate cancer, smoking status, education, physical activity, and physician visit over the past 12 months | 1.43 (1.01-2.03) |
| Genkinger et al., 2020 [2] | Diet and Cancer Pooling Project (DCPP) | World-wide pooling project (Mean age at | 1365/586,361 | Per 10 cm increase | 1.07 | 1.01-1.13 | Age, year of questionnaire return, race, education, marital status, alcohol, smoking | 1.07 (1.01-1.13) |

| | | | | | | | | |
|---|---|---|---|---|---|---|---|---|
| | | baseline: not available<br>Mean follow-up: not available) | | | | | habits, height, physical activity, prostate cancer family history, personal history of diabetes, multiple vitamin use, dietary calcium, dietary lycopene, and total energy intake | |
| Jochems et al., 2021 [22] | Five Swedish cohorts pooling project | Sweden (Mean age at baseline: 37.5y<br>Mean follow-up: 28y) | 387/58,457<br>(170 deaths from MDCS and 54 deaths from VIP likely to also be included in EPIC) | Per 10 cm increase | 1.07 | 0.95-1.20 | Stratified for cohort and birth period, and adjustment for age at study enrolment, smoking status at study enrolment, healthcare region, country of birth, highest education at study enrolment, and height | 1.07 (0.95-1.20) |
| Perez-Cornago et al., [this report] | UK Biobank | UK (Mean age at baseline: 56.5y<br>Mean follow-up: 10.9y) | 571/218,218 | Per 10 cm increase | 1.09 | 1.01-1.17 | Region, age at recruitment, Townsend deprivation score, ethnicity, lives with a wife or partner, smoking, physical activity, alcohol consumption, height, and diabetes | 1.09 (1.01-1.17) |

[1] Conversion explained in the Supplementary methods.

[2] Studies included: CPS II, Cancer Prevention Study II Nutrition Cohort; COSM, Cohort of Swedish Men; EPIC European Prospective Investigation into Cancer and Nutrition (follow-up only from baseline to 2004); HPFS, Health Professionals Follow-up Study; MCCS, Melbourne Collaborative Cohort Study; **MDCS,** Malmö Diet and Cancer Study; NIH-AARP, The NIH-AARP Diet and Health Study; VIP, Västerbotten Intervention Project**.**

Abbreviations: CI, confidence interval; HR, hazard ratio; Pca, prostate cancer.

**Supplementary table 3**. Characteristics of prospective studies and previous individual participant data meta-analysis of waist to hip ratio and prostate cancer death.

| Authors, Year (Reference) | Study | Country (Age, follow-up) | Pca death/total sample | Exposure categories | HR | 95% CI | Covariates | Conversion of HR (95%) to per 0.05 increment [1] |
|---|---|---|---|---|---|---|---|---|
| Martin et al., 2009 [10] | Nord-Trøndelag Health Study (HUNT 2) | Norway (Mean age at baseline: 48y<br>Mean follow-up: 9.3y) | 110/29,364 | Per 1 SD increase, 0.06 unit | 0.91 | 0.75-1.11 | Age, height, smoking, marital status, education, physical activity, International Prostate Symptom Score | 0.92 (0.79-1.09) |
| Perez-Cornago et al., 2017 [19] | European Prospective Investigation into Cancer and Nutrition (EPIC) | Europe (Mean age at baseline: 52y<br>Mean follow-up: 6y) | 870/141,896 | Per 0.1 unit increment | 1.18 | 1.08 - 1.28 | Age, education level, smoking status, marital status, diabetes, and physical activity | 1.07 (1.00-1.14) |
| Perez-Cornago et al., [this report] | UK Biobank | UK (Mean age at baseline: 56.5y<br>Mean follow-up: 10.9y) | 521340 | Per 0.05 unit increment | 1.10 | 1.03-1.17 | Region, age at recruitment, Townsend deprivation score, ethnicity, lives with a wife or partner, smoking, physical activity, alcohol consumption, height, and diabetes | 1.10 (1.03-1.17) |

[1] Conversion explained in the Supplementary methods.

Abbreviations: CI, confidence interval; HR, hazard ratio; Pca, prostate cancer.

**Supplementary table 4.** Baseline characteristics of participants according to fourths of BMI at recruitment in men from UK Biobank.

| | BMI | | | |
|---|---|---|---|---|
| **Characteristic** | **Q1** | **Q2** | **Q3** | **Q4** |
| No. of men | 55669 | 54197 | 54695 | 53286 |
| Sociodemographic | | | | |
| Age at recruitment (years), mean (SD) | 56.0 (8.4) | 56.7 (8.3) | 56.8 (8.2) | 56.7 (8.0) |
| Most deprived quintile, % (n) | 11644 (20.9) | 9671 (17.8) | 10449 (19.1) | 12876 (24.2) |
| No qualifications, % (n) | 6739 (12.1) | 7160 (13.2) | 7699 (14.1) | 7806 (14.6) |
| Black ethnicity, % (n) | 666 (1.2) | 744 (1.4) | 893 (1.6) | 915 (1.7) |
| Not in paid/self-employment, % (n) | 21148 (38.0) | 20529 (37.9) | 21025 (38.4) | 21594 (40.5) |
| Living with partner, % (n) | 40892 (73.5) | 42621 (78.6) | 43047 (78.7) | 39614 (74.3) |
| Anthropometric | | | | |
| Height (cm), mean (SD) | 176.2 (7.0) | 175.8 (6.8) | 175.5 (6.8) | 175.1 (6.8) |
| BMI (kg/m$^2$), mean (SD) | 23.2 (1.5) | 26.2 (0.7) | 28.6 (0.8) | 33.6 (3.4) |
| Body fat (%), mean (SD) | 19.5 (4.5) | 23.8 (3.6) | 26.6 (3.4) | 31.4 (4.1) |
| Waist circumference (cm), mean (SD) | 85.8 (6.3) | 93.1 (5.4) | 99.0 (5.7) | 110.4 (9.6) |
| Waist to hip ratio, mean (SD) | 0.886 (0.054) | 0.922 (0.050) | 0.948 (0.051) | 0.988 (0.059) |
| Plumper body size at age 10, % (n) | 3488 (6.3) | 5194 (9.6) | 7614 (13.9) | 12818 (24.1) |
| Lifestyle | | | | |
| Current cigarette smokers, % (n) | 8453 (15.2) | 6349 (11.7) | 6317 (11.5) | 6041 (11.3) |
| Drinking alcohol ≥ 20 g/day, % (n) | 21116 (37.9) | 24016 (44.3) | 25368 (46.4) | 23791 (44.6) |
| Physically inactive, % (n) | 12975 (23.3) | 13380 (24.7) | 15126 (27.7) | 18537 (34.8) |
| Health status | | | | |
| Vasectomy, % (n) | 2595 (4.7) | 2876 (5.3) | 3087 (5.6) | 2779 (5.2) |
| Hypertension, % (n) | 22203 (39.9) | 27266 (50.3) | 31023 (56.7) | 33158 (62.2) |
| Diabetes, % (n) | 1655 (3.0) | 2239 (4.1) | 3608 (6.6) | 7503 (14.1) |
| Prostate specific factors prior recruitment | | | | |
| PSA test before baseline, % (n) | 15253 (27.4) | 15731 (29.0) | 15466 (28.3) | 13897 (26.1) |
| Enlarged prostate, % (n) | 1803 (3.2) | 1896 (3.5) | 1805 (3.3) | 1558 (2.9) |
| Family history of prostate cancer, % (n) | 4208 (7.6) | 4159 (7.7) | 4121 (7.5) | 3868 (7.3) |

[1] Values are means (SD).

Abbreviations: BMI, body mass index; PSA, prostate specific antigen.

**Supplementary table 5.** Baseline characteristics of participants according to fourths of waist at recruitment in men from UK Biobank.

| | Waist | | | |
|---|---|---|---|---|
| **Characteristic** | **Q1** | **Q2** | **Q3** | **Q4** |
| No. of men | 56238 | 57568 | 50128 | 54291 |
| Sociodemographic | | | | |
| Age at recruitment (years), mean (SD) | 55.1 (8.5) | 56.4 (8.2) | 57.2 (8.0) | 57.7 (7.8) |
| Most deprived quintile, % (n) | 11779 (20.9) | 10546 (18.3) | 9584 (19.1) | 12892 (23.7) |
| No qualifications, % (n) | 7028 (12.5) | 7544 (13.1) | 7085 (14.1) | 7807 (14.4) |
| Black ethnicity, % (n) | 1077 (1.9) | 878 (1.5) | 667 (1.3) | 603 (1.1) |
| Not in paid/self-employment, % (n) | 19416 (34.5) | 21321 (37.0) | 19874 (39.6) | 23954 (44.1) |
| Living with partner, % (n) | 41577 (73.9) | 45268 (78.6) | 39378 (78.6) | 40154 (74.0) |
| Anthropometric | | | | |
| Height (cm), mean (SD) | 174.4 (6.8) | 175.4 (6.7) | 176.1 (6.8) | 176.7 (6.9) |
| BMI (kg/m$^2$), mean (SD) | 23.8 (2.1) | 26.4 (1.9) | 28.5 (2.1) | 32.9 (4.0) |
| Body fat (%), mean (SD) | 19.6 (4.5) | 23.9 (3.6) | 26.7 (3.4) | 31.2 (4.1) |
| Waist circumference (cm), mean (SD) | 83.9 (4.5) | 93.1 (2.0) | 99.8 (2.0) | 111.9 (8.1) |
| Waist to hip ratio, mean (SD) | 0.869 (0.045) | 0.921 (0.036) | 0.954 (0.038) | 1.003 (0.052) |
| Plumper body size at age 10, % (n) | 4076 (7.2) | 5925 (10.3) | 6820 (13.6) | 12370 (22.8) |
| Lifestyle | | | | |
| Current cigarette smokers, % (n) | 7885 (14.0) | 6891 (12.0) | 6024 (12.0) | 6447 (11.9) |
| Drinking alcohol ≥ 20 g/day, % (n) | 21381 (38.0) | 25446 (44.2) | 23243 (46.4) | 24334 (44.8) |
| Physically inactive, % (n) | 11425 (20.3) | 14336 (24.9) | 14385 (28.7) | 20076 (37.0) |
| Health status | | | | |
| Vasectomy, % (n) | 2821 (5.0) | 3067 (5.3) | 2701 (5.4) | 2754 (5.1) |
| Hypertension, % (n) | 22688 (40.3) | 29308 (50.9) | 28344 (56.5) | 33524 (61.7) |
| Diabetes, % (n) | 1490 (2.6) | 2388 (4.1) | 3304 (6.6) | 7905 (14.6) |
| Prostate specific factors prior recruitment | | | | |
| PSA test, % (n) | 14500 (25.8) | 16166 (28.1) | 14536 (29.0) | 15237 (28.1) |
| Enlarged prostate, % (n) | 1619 (2.9) | 1950 (3.4) | 1708 (3.4) | 1795 (3.3) |
| Family history of prostate cancer, % (n) | 4152 (7.4) | 4292 (7.5) | 3874 (7.7) | 4063 (7.5) |

[1] Values are means (SD).

Abbreviations: BMI, body mass index; PSA, prostate specific antigen.

**Supplementary table 6:** Mean and SD in men from UK Biobank with available imaging data (up to 4,800 men).

| **Imaging data** | **Mean (SD)** |
|---|---|
| MRI measures | |
| Total adipose tissue volume, L | 19.64 (6.48) |
| Visceral adipose tissue, L | 4.88 (2.32) |
| Abdominal subcutaneous adipose tissue volume, L | 5.91 (2.53) |
| Muscle fat infiltration, % | 6.82 (1.71) |
| Liver proton density fat fraction, % | 4.71 (4.72) |
| DXA measures | |
| Total tissue fat percentage, % | 30.29 (6.49) |
| Trunk fat mass, g | 42.74 (8.05) |
| Trunk fat percentage, % | 35.53 (8.59) |
| Android fat mass, g | 2,711 (1,247) |
| Android fat percentage, % | 38.06 (10.06) |
| Gynoid fat mass | 3,600 (1,327) |
| Gynoid fat % | 28.80 (6.05) |
| Visceral adipose tissue mass, g | 1,697 (952) |
| Visceral adipose tissue volume, $cm^3$ | 1,798 (1,009) |

**Supplementary table 7.** Pearson correlation coefficients between main adiposity measurements at baseline in 218,246 men from UK Biobank.

| | **BMI** | **Body fat %** | **Waist circumference** | **Waist to hip ratio** |
|---|---|---|---|---|
| BMI | 1.000 | | | |
| Body fat % | 0.799 | 1.000 | | |
| Waist circumference | 0.878 | 0.793 | 1.000 | |
| Waist to hip ratio | 0.593 | 0.625 | 0.793 | 1.000 |

All P values were < 0.001

**Supplementary table 8.** Pearson correlation coefficients between adiposity measurements (**imaging visit**) with MRI adiposity measurements from the imagining in up to 4,642 men from UK Biobank.

| MRI measurements | BMI | Body fat percentage | Waist | WHR | Total adipose tissue volume | Visceral adipose tissue | Abdominal subcutaneous adipose tissue volume | Muscle mass infiltration | Liver proton density fat fraction |
|---|---|---|---|---|---|---|---|---|---|
| BMI | 1.000 | | | | | | | | |
| Body fat percentage | 0.777 | 1.000 | | | | | | | |
| Waist | 0.861 | 0.773 | 1.000 | | | | | | |
| WHR | 0.574 | 0.588 | 0.777 | 1.000 | | | | | |
| Total adipose tissue volume | 0.876 | 0.841 | 0.886 | 0.604 | 1.000 | | | | |
| Visceral adipose tissue | 0.779 | 0.763 | 0.789 | 0.650 | 0.853 | 1.000 | | | |
| Abdominal subcutaneous adipose tissue volume | 0.852 | 0.788 | 0.849 | 0.552 | 0.940 | 0.694 | 1.000 | | |
| Muscle mass infiltration | 0.442 | 0.540 | 0.485 | 0.416 | 0.551 | 0.509 | 0.430 | 1.000 | |
| Liver proton density fat fraction | 0.430 | 0.414 | 0.412 | 0.363 | 0.430 | 0.526 | 0.358 | 0.212 | 1.000 |

All P values were < 0.001

Abbreviations: BMI, body mass index; MRI, magnetic resonance imaging; WHR, waist to hip ratio.

**Supplementary table 9.** Pearson correlation coefficients between adiposity measurements (**imaging visit**) with DXA adiposity measurements from the imagining in up to 4,642 men from UK Biobank.

| DXA measurements | BMI | Body fat percentage | Waist | WHR | Total tissue fat % | Trunk fat mass | Truck fat % | Android fat mass | Android fat % | Gynoid fat mass | Gynoid fat % | VAT mass | VAT % |
|---|---|---|---|---|---|---|---|---|---|---|---|---|---|
| BMI | 1.000 | | | | | | | | | | | | |
| Body fat percentage | 0.777 | 1.000 | | | | | | | | | | | |
| Waist | 0.861 | 0.773 | 1.000 | | | | | | | | | | |
| WHR | 0.574 | 0.588 | 0.777 | 1.000 | | | | | | | | | |
| Total tissue fat % | 0.761 | 0.885 | 0.781 | 0.625 | 1.000 | | | | | | | | |
| Trunk fat mass | 0.891 | 0.709 | 0.885 | 0.578 | 0.718 | 1.000 | | | | | | | |
| Truck fat % | 0.765 | 0.864 | 0.780 | 0.650 | 0.979 | 0.731 | 1.000 | | | | | | |
| Android fat mass | 0.889 | 0.834 | 0.889 | 0.660 | 0.904 | 0.909 | 0.915 | 1.000 | | | | | |
| Android fat % | 0.745 | 0.847 | 0.757 | 0.643 | 0.962 | 0.710 | 0.993 | 0.902 | 1.000 | | | | |
| Gynoid fat mass | 0.853 | 0.768 | 0.846 | 0.496 | 0.830 | 0.857 | 0.775 | 0.878 | 0.737 | 1.000 | | | |
| Gynoid fat % | 0.701 | 0.809 | 0.729 | 0.523 | 0.917 | 0.635 | 0.845 | 0.782 | 0.805 | 0.881 | 1.000 | | |
| VAT mass | 0.809 | 0.761 | 0.808 | 0.658 | 0.820 | 0.842 | 0.860 | 0.936 | 0.850 | 0.738 | 0.647 | 1.000 | |
| VAT % | 0.809 | 0.761 | 0.808 | 0.658 | 0.820 | 0.842 | 0.860 | 0.936 | 0.850 | 0.738 | 0.647 | 1.000 | 1.000 |

All P values were < 0.001

Abbreviations: BMI, body mass index; DXA, Dual-energy X-ray absorptiometry; VAT, visceral adipose tissue; WHR, waist to hip ratio.

**Supplementary table 10**. Geometric means of selected MRI measurements by tenths of anthropometric measurements **at the imaging visit** in men from UK Biobank.

| Anthropometric measurements at imaging visit | | Total adipose tissue volume | | Visceral adipose tissue | | Abdominal subcutaneous adipose tissue volume | | Muscle mass infiltration | | Liver proton density fat fraction | |
|---|---|---|---|---|---|---|---|---|---|---|---|
| | **Tenths** | **n** | **Mean (min-max)** | **n** | **Mean (min-max)** | **n** | **Mean (min-max)** | **n** | **Mean (min-max)** | **n** | **Mean (min-max)** |
| **BMI** | 1 | 386 | 11.25 (10.95-11.55) | 455 | 1.97 (1.84-2.10) | 455 | 2.98 (2.85-3.11) | 436 | 5.74 (5.61-5.87) | 448 | 1.76 (0.65-9.47) |
| | 2 | 386 | 13.93 (13.63-14.23) | 461 | 2.92 (2.79-3.05) | 461 | 3.94 (3.81-4.07) | 448 | 6.02 (5.89-6.15) | 455 | 2.55 (0.73-13.20) |
| | 3 | 389 | 15.65 (15.35-15.94) | 460 | 3.51 (3.38-3.64) | 460 | 4.44 (4.31-4.57) | 451 | 6.24 (6.12-6.37) | 453 | 2.96 (0.80-25.19) |
| | 4 | 402 | 17.01 (16.72-17.31) | 478 | 4.03 (3.90-4.15) | 479 | 4.87 (4.74-5.00) | 462 | 6.53 (6.40-6.66) | 478 | 3.47 (0.89-21.67) |
| | 5 | 380 | 18.50 (18.20-18.80) | 464 | 4.57 (4.44-4.70) | 464 | 5.30 (5.17-5.43) | 450 | 6.68 (6.55-6.81) | 461 | 4.05 (0.67-30.18) |
| | 6 | 408 | 19.76 (19.47-20.05) | 479 | 4.97 (4.84-5.10) | 479 | 5.77 (5.64-5.89) | 465 | 6.72 (6.60-6.85) | 475 | 4.51 (0.86-27.21) |
| | 7 | 382 | 21.64 (21.34-21.95) | 478 | 5.57 (5.44-5.70) | 478 | 6.37 (6.25-6.50) | 456 | 7.07 (6.94-7.20) | 472 | 5.10 (0.86-26.08) |
| | 8 | 384 | 23.43 (23.13-23.74) | 469 | 6.08 (5.95-6.21) | 469 | 6.93 (6.80-7.06) | 454 | 7.24 (7.11-7.37) | 463 | 6.39 (0.85-32.50) |
| | 9 | 347 | 26.38 (26.07-26.70) | 450 | 6.90 (6.76-7.03) | 450 | 8.05 (7.92-8.18) | 424 | 7.60 (7.47-7.74) | 443 | 6.90 (0.89-28.15) |
| | 10 | 298 | 32.40 (32.05-32.74) | 436 | 8.48 (8.35-8.62) | 436 | 10.65 (10.52-10.79) | 408 | 8.44 (8.30-8.58) | 438 | 9.57 (1.01-34.04) |
| **Body fat percentage** | | | | | | | | | | | |
| | 1 | 375 | 10.88 (10.55-11.21) | 487 | 1.94 (1.81-2.08) | 487 | 2.93 (2.80-3.06) | 469 | 5.58 (5.46-5.70) | 192 | 1.47 (0.86-2.08) |
| | 2 | 379 | 14.02 (13.70-14.35) | 441 | 2.93 (2.80-3.07) | 441 | 3.92 (3.79-4.06) | 423 | 6.00 (5.87-6.13) | 215 | 2.20 (1.63-2.78) |
| | 3 | 416 | 16.09 (15.78-16.40) | 496 | 3.75 (3.62-3.88) | 497 | 4.61 (4.48-4.74) | 480 | 6.19 (6.07-6.31) | 213 | 2.91 (2.34-3.49) |
| | 4 | 383 | 16.94 (16.61-17.26) | 448 | 4.00 (3.87-4.14) | 448 | 4.89 (4.76-5.03) | 439 | 6.44 (6.32-6.57) | 199 | 2.97 (2.38-3.57) |
| | 5 | 415 | 18.74 (18.43-19.05) | 502 | 4.74 (4.62-4.87) | 502 | 5.48 (5.35-5.61) | 487 | 6.59 (6.47-6.71) | 237 | 4.00 (3.46-4.54) |
| | 6 | 356 | 20.38 (20.04-20.71) | 433 | 5.14 (5.00-5.28) | 433 | 5.99 (5.85-6.13) | 417 | 6.98 (6.85-7.11) | 195 | 4.24 (3.64-4.84) |
| | 7 | 343 | 21.17 (20.83-21.51) | 419 | 5.51 (5.36-5.65) | 419 | 6.26 (6.12-6.40) | 405 | 6.89 (6.76-7.02) | 199 | 5.48 (4.88-6.07) |
| | 8 | 392 | 23.56 (23.24-23.88) | 493 | 6.29 (6.16-6.42) | 493 | 7.13 (7.00-7.26) | 472 | 7.33 (7.21-7.45) | 214 | 6.12 (5.55-6.69) |
| | 9 | 353 | 26.09 (25.75-26.43) | 443 | 6.82 (6.68-6.96) | 443 | 7.91 (7.77-8.05) | 418 | 7.74 (7.62-7.87) | 219 | 6.59 (6.03-7.16) |
| | 10 | 299 | 31.87 (31.50-32.24) | 412 | 8.25 (8.10-8.39) | 412 | 10.66 (10.51-10.80) | 391 | 8.71 (8.58-8.84) | 187 | 8.55 (7.93-9.16) |

All models are adjusted for sex and height.

Abbreviations: BMI, body mass index; WHR, waist to hip ratio.

**Supplementary table 10**. Continued

| Anthropometric measurements at imaging visit | | Total adipose tissue volume | | Visceral adipose tissue | | Abdominal subcutaneous adipose tissue volume | | Muscle mass infiltration | | Liver proton density fat fraction | |
|---|---|---|---|---|---|---|---|---|---|---|---|
| | Tenths | n | Mean (min-max) | n | Mean (min-max) | n | Mean (min-max) | n | Mean (min-max) | n | Mean (min-max) |
| **Waist circumference** | | | | | | | | | | | |
| | 1 | 476 | 11.66 (11.38-11.94) | 560 | 2.11 (1.99-2.23) | 560 | 3.11 (2.99-3.22) | 540 | 5.70 (5.58-5.82) | 554 | 1.99 (0.70-19.09) |
| | 2 | 460 | 14.58 (14.29-14.86) | 552 | 3.11 (2.99-3.23) | 552 | 4.10 (3.98-4.22) | 541 | 6.06 (5.95-6.18) | 545 | 2.74 (0.65-18.67) |
| | 3 | 329 | 16.30 (15.97-16.64) | 372 | 3.71 (3.56-3.85) | 372 | 4.62 (4.48-4.76) | 369 | 6.36 (6.22-6.50) | 369 | 3.26 (0.75-18.77) |
| | 4 | 513 | 17.49 (17.22-17.76) | 615 | 4.15 (4.04-4.26) | 616 | 5.02 (4.91-5.13) | 600 | 6.56 (6.45-6.67) | 612 | 3.67 (0.74-24.33) |
| | 5 | 341 | 19.05 (18.72-19.38) | 408 | 4.66 (4.52-4.80) | 408 | 5.50 (5.37-5.64) | 393 | 6.68 (6.54-6.81) | 406 | 4.46 (0.85-25.19) |
| | 6 | 462 | 20.87 (20.59-21.16) | 567 | 5.36 (5.24-5.48) | 567 | 6.01 (5.90-6.12) | 547 | 7.00 (6.89-7.12) | 558 | 4.95 (0.86-30.18) |
| | 7 | 271 | 22.03 (21.66-22.40) | 328 | 5.77 (5.61-5.93) | 328 | 6.40 (6.25-6.56) | 321 | 7.14 (6.99-7.29) | 324 | 5.30 (1.00-32.50) |
| | 8 | 386 | 24.07 (23.76-24.38) | 492 | 6.31 (6.18-6.43) | 492 | 7.23 (7.10-7.35) | 471 | 7.26 (7.14-7.39) | 486 | 6.11 (0.86-28.39) |
| | 9 | 304 | 27.77 (27.41-28.12) | 400 | 7.24 (7.10-7.38) | 400 | 8.60 (8.47-8.74) | 368 | 7.81 (7.66-7.95) | 394 | 7.56 (0.89-27.63) |
| | 10 | 220 | 33.55 (33.14-33.96) | 336 | 8.73 (8.57-8.88) | 336 | 11.23 (11.08-11.38) | 304 | 8.75 (8.60-8.91) | 338 | 9.75 (1.08-34.04) |
| **WHR** | | | | | | | | | | | |
| | 1 | 427 | 13.24 (12.76-13.73) | 515 | 2.44 (2.29-2.59) | 515 | 3.71 (3.53-3.89) | 491 | 5.82 (5.69-5.95) | 509 | 2.17 (0.73-19.09) |
| | 2 | 440 | 15.86 (15.39-16.33) | 540 | 3.42 (3.27-3.57) | 541 | 4.51 (4.34-4.69) | 523 | 6.30 (6.18-6.42) | 539 | 2.88 (0.65-21.60) |
| | 3 | 449 | 16.97 (16.50-17.44) | 538 | 3.88 (3.74-4.03) | 538 | 4.99 (4.81-5.16) | 530 | 6.35 (6.23-6.47) | 532 | 3.52 (0.78-24.33) |
| | 4 | 445 | 18.36 (17.89-18.83) | 539 | 4.38 (4.23-4.53) | 539 | 5.40 (5.23-5.58) | 520 | 6.56 (6.43-6.68) | 530 | 4.12 (0.67-28.07) |
| | 5 | 403 | 19.77 (19.28-20.27) | 487 | 4.88 (4.72-5.03) | 487 | 5.83 (5.65-6.02) | 465 | 6.85 (6.72-6.98) | 483 | 4.58 (0.80-30.18) |
| | 6 | 378 | 20.62 (20.11-21.13) | 463 | 5.25 (5.09-5.41) | 463 | 6.17 (5.98-6.35) | 447 | 6.85 (6.72-6.99) | 459 | 4.77 (0.93-25.47) |
| | 7 | 382 | 22.14 (21.64-22.65) | 460 | 5.79 (5.63-5.95) | 460 | 6.69 (6.50-6.88) | 444 | 7.05 (6.91-7.18) | 455 | 5.73 (0.75-28.39) |
| | 8 | 352 | 23.53 (23.00-24.06) | 439 | 6.26 (6.10-6.42) | 439 | 7.19 (6.99-7.38) | 420 | 7.44 (7.30-7.57) | 430 | 6.49 (0.86-32.50) |
| | 9 | 276 | 25.48 (24.89-26.08) | 369 | 6.92 (6.74-7.09) | 369 | 7.91 (7.70-8.12) | 348 | 7.82 (7.67-7.97) | 367 | 7.12 (1.07-29.43) |
| | 10 | 210 | 27.87 (27.19-28.56) | 280 | 8.12 (7.92-8.33) | 280 | 9.07 (8.83-9.31) | 266 | 8.24 (8.07-8.42) | 282 | 8.73 (1.16-34.04) |

All models are adjusted for sex and height.

Abbreviations: BMI, body mass index; WHR, waist to hip ratio.

**Supplementary table 11**. Geometric means of selected DXA measurements by tenths of anthropometric measurements **at the imaging visit** in men from UK Biobank.

| Anthropometric measurements at imaging visit | | Total tissue fat percentage | | Trunk fat mass | | Truck fat percentage | | Visceral adipose tissue mass | | Android fat mass | | Android fat percentage | |
|---|---|---|---|---|---|---|---|---|---|---|---|---|---|
| | Tenths | n | Mean (min-max) | n | Mean (min-max) | n | Mean (min-max) | n | Mean (min-max) | n | Mean (min-max) | n | Mean (min-max) |
| **BMI** | | | | | | | | | | | | | |
| | 1 | 216 | 21.2 (20.6-21.8) | 216 | 32.1 (31.7-32.4) | 216 | 22.4 (21.7-23.1) | 215 | 554 (479-630) | 216 | 1,077 (992-1,161) | 216 | 1,077 (992-1,161) |
| | 2 | 229 | 24.7 (24.1-25.2) | 229 | 35.8 (35.5-36.2) | 229 | 28.0 (27.3-28.7) | 228 | 871 (798-944) | 229 | 1,617 (1,535-1,700) | 229 | 1,617 (1,535-1,700) |
| | 3 | 223 | 26.9 (26.3-27.4) | 223 | 37.6 (37.2-38.0) | 223 | 30.9 (30.2-31.6) | 222 | 1,101 (1,027-1,175) | 223 | 1,920 (1,837-2,002) | 223 | 1,920 (1,837-2,002) |
| | 4 | 226 | 28.2 (27.6-28.7) | 226 | 39.2 (38.8-39.6) | 226 | 32.9 (32.2-33.6) | 225 | 1,271 (1,197-1,344) | 226 | 2,163 (2,080-2,245) | 226 | 2,163 (2,080-2,245) |
| | 5 | 239 | 29.6 (29.1-30.2) | 239 | 40.8 (40.5-41.2) | 239 | 35.0 (34.3-35.6) | 238 | 1,481 (1,410-1,553) | 239 | 2,419 (2,339-2,499) | 239 | 2,419 (2,339-2,499) |
| | 6 | 253 | 30.7 (30.2-31.2) | 253 | 42.6 (42.2-43.0) | 253 | 36.5 (35.9-37.2) | 250 | 1,716 (1,647-1,786) | 253 | 2,688 (2,610-2,766) | 253 | 2,688 (2,610-2,766) |
| | 7 | 258 | 32.1 (31.6-32.7) | 258 | 44.3 (44.0-44.7) | 258 | 38.2 (37.6-38.9) | 255 | 1,901 (1,832-1,970) | 258 | 2,951 (2,873-3,028) | 258 | 2,951 (2,873-3,028) |
| | 8 | 251 | 33.4 (32.8-33.9) | 251 | 46.4 (46.0-46.8) | 251 | 39.9 (39.2-40.6) | 250 | 2,154 (2,085-2,224) | 251 | 3,279 (3,201-3,357) | 251 | 3,279 (3,201-3,357) |
| | 9 | 237 | 35.0 (34.4-35.5) | 237 | 49.2 (48.8-49.5) | 237 | 42.0 (41.3-42.7) | 236 | 2,454 (2,382-2,525) | 237 | 3,684 (3,603-3,764) | 237 | 3,684 (3,603-3,764) |
| | 10 | 227 | 39.6 (39.0-40.1) | 227 | 57.9 (57.5-58.3) | 227 | 47.3 (46.6-48.0) | 215 | 3,354 (3,278-3,429) | 227 | 5,065 (4,983-5,147) | 227 | 5,065 (4,983-5,147) |
| **Body fat %** | | | | | | | | | | | | | |
| | 1 | 203 | 19.5 (19.0-19.9) | 203 | 33.5 (32.9-34.2) | 203 | 20.6 (20.0-21.2) | 201 | 520 (436-604) | 203 | 1,000 (908-1,093) | 203 | 1,000 (908-1,093) |
| | 2 | 225 | 23.7 (23.3-24.2) | 225 | 36.5 (35.9-37.1) | 225 | 26.9 (26.3-27.5) | 223 | 870 (790-949) | 225 | 1,573 (1,485-1,660) | 225 | 1,573 (1,485-1,660) |
| | 3 | 246 | 26.5 (26.1-26.9) | 246 | 38.8 (38.2-39.4) | 246 | 30.9 (30.4-31.5) | 244 | 1,174 (1,098-1,251) | 246 | 1,995 (1,911-2,079) | 246 | 1,995 (1,911-2,079) |
| | 4 | 218 | 27.8 (27.4-28.2) | 218 | 39.5 (38.9-40.1) | 218 | 32.5 (31.9-33.1) | 217 | 1,276 (1,195-1,357) | 218 | 2,150 (2,061-2,239) | 218 | 2,150 (2,061-2,239) |
| | 5 | 255 | 29.4 (29.0-29.8) | 255 | 41.2 (40.6-41.7) | 255 | 34.8 (34.2-35.3) | 255 | 1,498 (1,423-1,572) | 255 | 2,439 (2,357-2,522) | 255 | 2,439 (2,357-2,522) |
| | 6 | 226 | 31.3 (30.9-31.7) | 226 | 42.9 (42.3-43.5) | 226 | 37.2 (36.6-37.8) | 226 | 1,754 (1,675-1,833) | 226 | 2,781 (2,694-2,868) | 226 | 2,781 (2,694-2,868) |
| | 7 | 232 | 32.4 (32.0-32.8) | 232 | 43.6 (43.0-44.1) | 232 | 38.5 (38.0-39.1) | 229 | 1,872 (1,794-1,951) | 232 | 2,944 (2,858-3,031) | 232 | 2,944 (2,858-3,031) |
| | 8 | 251 | 34.1 (33.7-34.5) | 251 | 46.1 (45.5-46.6) | 251 | 40.8 (40.3-41.4) | 249 | 2,218 (2,142-2,293) | 251 | 3,341 (3,258-3,424) | 251 | 3,341 (3,258-3,424) |
| | 9 | 254 | 35.9 (35.5-36.3) | 254 | 48.0 (47.5-48.6) | 254 | 42.8 (42.2-43.3) | 251 | 2,432 (2,357-2,508) | 254 | 3,690 (3,607-3,772) | 254 | 3,690 (3,607-3,772) |
| | 10 | 219 | 40.4 (40.0-40.8) | 219 | 56.0 (55.4-56.7) | 219 | 47.8 (47.2-48.4) | 209 | 3,189 (3,106-3,271) | 219 | 4,934 (4,845-5,023) | 219 | 4,934 (4,845-5,023) |

**Supplementary table 11**. Continued.

| Anthropometric measurements at imaging visit | Tenths | Total tissue fat percentage n | Mean (min-max) | Trunk fat mass n | Mean (min-max) | Truck fat percentage n | Mean (min-max) | Visceral adipose tissue mass n | Mean (min-max) | Android fat mass n | Mean (min-max) | Android fat percentage n | Mean (min-max) |
|---|---|---|---|---|---|---|---|---|---|---|---|---|---|
| Anthropometric measurements at imaging visit | | | | | | | | | | | | | |
| **Waist circumference** | | | | | | | | | | | | | |
| | 1 | 268 | 21.2 (20.7-21.7) | 268 | 33.4 (33.0-33.9) | 268 | 22.7 (22.0-23.3) | 266 | 595 (526-665) | 268 | 1,156 (1,079-1,233) | 268 | 1,156 (1,079-1,233) |
| | 2 | 253 | 25.2 (24.7-25.7) | 253 | 36.6 (36.1-37.1) | 253 | 28.8 (28.2-29.4) | 252 | 962 (891-1,033) | 253 | 1,713 (1,635-1,792) | 253 | 1,713 (1,635-1,792) |
| | 3 | 199 | 27.1 (26.6-27.7) | 199 | 38.4 (37.9-39.0) | 199 | 31.4 (30.7-32.1) | 198 | 1,163 (1,083-1,243) | 199 | 1,991 (1,903-2,080) | 199 | 1,991 (1,903-2,080) |
| | 4 | 323 | 28.7 (28.2-29.1) | 323 | 40.0 (39.6-40.4) | 323 | 33.5 (32.9-34.0) | 321 | 1,347 (1,284-1,410) | 323 | 2,257 (2,188-2,326) | 323 | 2,257 (2,188-2,326) |
| | 5 | 208 | 29.8 (29.3-30.3) | 208 | 41.6 (41.1-42.1) | 208 | 35.3 (34.6-36.0) | 206 | 1,526 (1,448-1,604) | 208 | 2,496 (2,410-2,582) | 208 | 2,496 (2,410-2,582) |
| | 6 | 305 | 31.5 (31.0-31.9) | 305 | 43.2 (42.8-43.6) | 305 | 37.5 (36.9-38.0) | 304 | 1,823 (1,758-1,887) | 305 | 2,807 (2,736-2,879) | 305 | 2,807 (2,736-2,879) |
| | 7 | 162 | 32.4 (31.8-33.0) | 162 | 44.8 (44.2-45.4) | 162 | 38.8 (38.0-39.6) | 161 | 1,989 (1,900-2,077) | 162 | 3,050 (2,953-3,148) | 162 | 3,050 (2,953-3,148) |
| | 8 | 256 | 34.2 (33.7-34.7) | 256 | 46.8 (46.4-47.3) | 256 | 41.1 (40.5-41.7) | 254 | 2,268 (2,197-2,339) | 256 | 3,438 (3,360-3,516) | 256 | 3,438 (3,360-3,516) |
| | 9 | 213 | 36.8 (36.3-37.4) | 213 | 50.5 (50.0-51.0) | 213 | 44.1 (43.4-44.8) | 211 | 2,671 (2,593-2,749) | 213 | 3,994 (3,908-4,079) | 213 | 3,994 (3,908-4,079) |
| | 10 | 173 | 40.7 (40.1-41.3) | 173 | 58.9 (58.3-59.4) | 173 | 48.6 (47.8-49.4) | 162 | 3,481 (3,393-3,570) | 173 | 5,314 (5,219-5,409) | 173 | 5,314 (5,219-5,409) |
| **WHR** | | | | | | | | | | | | | |
| | 1 | 251 | 22.8 (22.2-23.4) | 251 | 35.4 (34.7-36.1) | 251 | 24.9 (24.1-25.8) | 249 | 748 (659-838) | 251 | 1,403 (1,288-1,518) | 251 | 1,403 (1,288-1,518) |
| | 2 | 267 | 26.1 (25.5-26.7) | 267 | 38.2 (37.5-38.9) | 267 | 29.7 (28.9-30.5) | 266 | 1,045 (959-1,131) | 267 | 1,858 (1,747-1,969) | 267 | 1,858 (1,747-1,969) |
| | 3 | 280 | 28.0 (27.4-28.6) | 280 | 39.7 (39.0-40.3) | 280 | 32.5 (31.8-33.3) | 279 | 1,284 (1,200-1,368) | 280 | 2,203 (2,094-2,311) | 280 | 2,203 (2,094-2,311) |
| | 4 | 282 | 29.4 (28.8-30.0) | 282 | 41.5 (40.8-42.2) | 282 | 34.5 (33.7-35.3) | 281 | 1,485 (1,401-1,568) | 282 | 2,474 (2,366-2,582) | 282 | 2,474 (2,366-2,582) |
| | 5 | 249 | 30.3 (29.6-30.9) | 249 | 42.5 (41.7-43.2) | 249 | 35.8 (35.0-36.6) | 249 | 1,673 (1,583-1,762) | 249 | 2,654 (2,538-2,769) | 249 | 2,654 (2,538-2,769) |
| | 6 | 237 | 31.4 (30.7-32.0) | 237 | 43.3 (42.5-44.0) | 237 | 37.1 (36.3-38.0) | 235 | 1,787 (1,695-1,878) | 237 | 2,816 (2,698-2,934) | 237 | 2,816 (2,698-2,934) |
| | 7 | 242 | 33.1 (32.5-33.8) | 242 | 45.4 (44.6-46.1) | 242 | 39.6 (38.8-40.4) | 240 | 2,110 (2,019-2,201) | 242 | 3,231 (3,114-3,348) | 242 | 3,231 (3,114-3,348) |
| | 8 | 227 | 34.0 (33.4-34.7) | 227 | 46.6 (45.8-47.4) | 227 | 40.5 (39.6-41.3) | 223 | 2,252 (2,158-2,346) | 227 | 3,416 (3,295-3,537) | 227 | 3,416 (3,295-3,537) |
| | 9 | 177 | 35.3 (34.5-36.0) | 177 | 48.6 (47.7-49.5) | 177 | 42.5 (41.5-43.4) | 173 | 2,520 (2,413-2,627) | 177 | 3,755 (3,618-3,892) | 177 | 3,755 (3,618-3,892) |
| | 10 | 148 | 38.0 (37.2-38.8) | 148 | 53.6 (52.6-54.5) | 148 | 45.7 (44.7-46.8) | 140 | 3,107 (2,988-3,226) | 148 | 4,565 (4,415-4,714) | 148 | 4,565 (4,415-4,714) |

All models are adjusted for sex and height.

Abbreviations: BMI, body mass index; WHR, waist to hip ratio.

**Supplementary Table 12.** Minimally- and multivariable-adjusted hazard ratios (95% CI) for prostate cancer death in relation to adiposity measurements at baseline in men from UK Biobank.

| Anthropometric | Total n | PCa deaths | HR (95% CI) Minimally-adjusted | HR (95% CI) Multivariable-adjusted | P trend |
|---|---|---|---|---|---|
| BMI, kg/m$^2$ | | | | | |
| Q1, <=25.0 | 55669 | 151 | 1 ref | 1 ref | |
| Q2, 25.1 - 27.2 | 54197 | 141 | 0.89 (0.71 - 1.12) | 0.90 (0.71 - 1.13) | |
| Q3, 27.3-30.0 | 54695 | 183 | 1.15 (0.92 - 1.42) | 1.15 (0.92 - 1.43) | |
| Q4, >=30.1 | 53286 | 156 | 1.05 (0.84 - 1.32) | 1.04 (0.83 - 1.31) | |
| Per 5 kg/m$^2$ increase | 217841 | 631 | 1.10 (1.00 - 1.21) | 1.10 (1.00 - 1.21) | 0.059 |
| Body fat, % | | | | | |
| Q1, <=21.5 | 56238 | 131 | 1 ref | 1 ref | |
| Q2, 21.6 - 25.4 | 57568 | 127 | 0.79 (0.62 - 1.01) | 0.81 (0.63 - 1.03) | |
| Q3, 25.5-29.1 | 50128 | 169 | 0.98 (0.78 - 1.23) | 1.00 (0.79 - 1.25) | |
| Q4, >=29.2 | 54291 | 190 | 1.00 (0.80 - 1.25) | 1.02 (0.81 - 1.29) | |
| Per 5 % increase | 214219 | 617 | 1.03 (0.96 - 1.10) | 1.03 (0.96 - 1.11) | 0.398 |
| Waist circumference, cm | | | | | |
| Q1, <=89 | 56238 | 119 | 1 ref | 1 ref | |
| Q2, 89.1 - 96.0 | 57568 | 154 | 1.10 (0.87 - 1.40) | 1.09 (0.86 - 1.39) | |
| Q3, 96.1-103.0 | 50128 | 158 | 1.22 (0.96 - 1.54) | 1.19 (0.94 - 1.52) | |
| Q4, >=103.1 | 54291 | 200 | 1.41 (1.12 - 1.77) | 1.36 (1.07 - 1.72) | |
| Per 10 cm increase | 218218 | 631 | 1.11 (1.03 - 1.19) | 1.09 (1.02 - 1.18) | 0.018 |
| Waist to hip ratio | | | | | |
| Q1, <=0.892 | 54636 | 114 | 1 ref | 1 ref | |
| Q2, 0.893 - 0.934 | 54486 | 138 | 1.03 (0.80 - 1.32) | 1.03 (0.80 - 1.32) | |
| Q3, 0.935-0.978 | 54568 | 176 | 1.18 (0.93 - 1.50) | 1.19 (0.93 - 1.51) | |
| Q4, >=0.979 | 54483 | 203 | 1.27 (1.01 - 1.60) | 1.28 (1.01 - 1.62) | |
| Per 0.05 increase | 218166 | 631 | 1.08 (1.02 - 1.15) | 1.09 (1.02 - 1.16) | 0.010 |

Cox regression analyses.

Minimally-adjusted models are stratified by region and age at recruitment and adjusted for age (underlying time variable).

Multivariable-adjusted models are stratified by region and age at recruitment and adjusted for age (underlying time variable), Townsend deprivation score, ethnicity, lives with a wife or partner, smoking, physical activity, alcohol consumption, height, and diabetes. Full details for each covariate are provided in the statistical section.

Abbreviations: BMI, body mass index; PCa, prostate cancer.

**Supplementary table 13.** Multivariable-adjusted hazard ratios (95 % CI) for prostate cancer in relation to BMI, waist circumference and WHR using the WHO cut-off points at recruitment in men from UK Biobank.

| | **BMI (kg/m$^2$)** | | | **Waist circumference (cm)** | | | **WHR** | |
|---|---|---|---|---|---|---|---|---|
| | **<25** | **25-29.9** | **≥30** | **<94** | **94-101.9** | **≥ 102 (higher risk)** | **<0.90** | **≥ 0.90** |
| N pca deaths | 147 | 313 | 171 | 215 | 174 | 242 | 133 | 498 |
| PCa death | 1 ref | 0.98 (0.81 - 1.20) | 1.06 (0.85 - 1.34) | 1 ref | 0.97 (0.79 - 1.18) | 1.22 (1.00 - 1.48) | 1 ref | 1.17 (0.96 - 1.43) |

Cox regression analysis. Multivariable-adjusted models are stratified by region and age at recruitment and adjusted for age (underlying time variable), Townsend deprivation score, ethnicity, lives with a wife or partner, smoking, physical activity, alcohol consumption, height, and diabetes . Full details for each covariate are provided in the statistical section.

Abbreviations: BMI, body mass index; PCa, prostate cancer; WHR, waist to hip ratio.
